# The aconitate decarboxylase 1/itaconate pathway modulates immune dysregulation and associates with cardiovascular disease markers in SLE

**DOI:** 10.1101/2024.02.20.24303097

**Authors:** Eduardo Patiño-Martinez, Shuichiro Nakabo, Kan Jiang, Carmelo Carmona-Rivera, Wanxia Li Tsai, Dillon Claybaugh, Zu-Xi Yu, Aracely Romero, Eric Bohrnsen, Benjamin Schwarz, Miguel A. Solís-Barbosa, Luz P. Blanco, Mohammad Naqi, Yenealem Temesgen-Oyelakim, Michael Davis, Zerai Manna, Nehal Mehta, Faiza Naz, Stephen Brooks, Stefania dell’Orso, Sarfaraz Hasni, Mariana J. Kaplan

**Author notes:** Correspondence and reprint requests: Mariana J. Kaplan, M.D., Systemic Autoimmunity Branch; NIAMS/National Institutes of Health 10 Center Drive; 12N248C, Bethesda, MD 20892. There are no conflicts of interest.

## Abstract

**What is already known on this topic**

Aconitate Decarboxylase 1 (ACOD1) is an enzyme involved in the synthesis of itaconate, a metabolite generated during the Krebs cycle.
Itaconate has been identified as an immunomodulatory molecule
ACOD1/Itaconate has been studied in the context of various inflammatory and autoimmune diseases, including sepsis, inflammatory bowel disease and rheumatoid arthritis. In these conditions, dysregulation of itaconate metabolism has been associated with altered immune responses and disease progression.

**What this study adds**

1. Upon stimulation with lupus-relevant stimuli, ACOD1 expression is induced in myeloid cells.
2. IN an induced mouse model of lupus, ACOD1 knockout (Acod1-/-) mice exhibit exacerbated lupus-like symptoms, implicating dysregulation of this pathway in the induction and severity of autoimmunity features.
3. Itaconate serum levels are decreased in SLE patients, compared to healthy individuals. This decrease is associated with specific perturbed cardiometabolic parameters and subclinical atherosclerosis, indicating that modulating dysregulation of the itaconate pathway could have therapeutic benefits in this disease.

**How this study might affect research, practice or policy**

Given its immunomodulatory effects, ACOD1/itaconate and its derivatives may have potential therapeutic benefit for the treatment of autoimmune diseases. They may also serve as putative biomarkers of cardiovascular risk in this disease.

**Objective:** The Krebs cycle enzyme Aconitate Decarboxylase 1 (ACOD1) mediates itaconate synthesis in myeloid cells.. Previously, we reported that administration of 4-octyl itaconate abrogated lupus phenotype in mice. Here, we explore the role of the endogenous ACOD1/itaconate pathway in the development of murine lupus as well as their relevance in premature cardiovascular damage in SLE.

**Methods:** We characterized Acod1 protein expression in bone marrow-derived macrophages and human monocyte-derived macrophages, following a TLR7 agonist (imiquimod, IMQ). Wild type and Acod1^-/-^ mice were exposed to topical IMQ for 5 weeks to induce an SLE phenotype and immune dysregulation was quantified. Itaconate serum levels were quantified in SLE patients and associated to cardiometabolic parameters and disease activity.

**Results:** ACOD1 was induced in mouse bone marrow-derived macrophages (BMDM) and human monocyte-derived macrophages following in vitro TLR7 stimulation. This induction was partially dependent on type I Interferon receptor signaling and specific intracellular pathways. In the IMQ-induced mouse model of lupus, ACOD1 knockout (*Acod1*^-/-^) displayed disruptions of the splenic architecture, increased serum anti-dsDNA and proinflammatory cytokine levels, enhanced kidney immune complex deposition and proteinuria, when compared to the IMQ-treated WT mice. Consistent with these results, *Acod1*^-/-^ BMDM exposed to IMQ showed higher proinflammatory features in vitro. Itaconate levels were decreased in SLE serum compared to healthy control sera, in association with specific perturbed cardiometabolic parameters and subclinical vascular disease.

**Conclusion:** These findings suggest that the ACOD1/itaconate pathway plays important immunomodulatory and vasculoprotective roles in SLE, supporting the potential therapeutic role of itaconate analogs in autoimmune diseases.

## INTRODUCTION

Systemic lupus erythematosus (SLE) is a chronic autoimmune disorder that can affect multiple organs and systems in the body. It is more prevalent in women and can present with a broad range of clinical symptoms and signs. The causes of SLE are complex and involve a combination of genetic, epigenetic and environmental factors that trigger a breakdown of immune tolerance. This leads to the activation of autoreactive lymphocytes, production of autoantibodies and innate immune dysregulation. The type I Interferon (IFN) pathway, myeloid aberrant responses (including enhanced synthesis of neutrophil extracellular traps (NETs)) and defective clearance of immune complexes and dead cells, all contribute to loss of tolerance, tissue damage and organ dysfunction in SLE (1, 2). Part of this dysregulation is driven by activation of intracellular sensors of nucleic acids, sch as Toll-like receptor-7 (TLR7). Furthermore, SLE patients have accelerated vascular damage that predisposes to premature heart attacks and stroke, aphenomenon considered to be driven by immune dysregulation (3, 4).

Several studies indicate that mitochondrial dysfunction, increased oxidative stress and other metabolic pathway alterations in both the innate and adaptive immune cells play a crucial role in the development and severity of SLE and its associated organ damage (5). Thus, pharmacological interventions targeting immunometabolism have emerged as potential approaches to managing SLE (6).

Aconitate decarboxylase 1 (ACOD1) is a mitochondrial enzyme that plays a vital role in cellular metabolism, oxidative stress and inflammatory responses (7). It mediates itaconate synthesis by decarboxylating cis-aconitate and can either promote or inhibit inflammation (8, 9). The ACOD1/itaconate axis is activated under stress conditions, including infections, and downstream of TLRs or cytokine stimulation in macrophages, monocytes and dendritic cells (DCs) (10).

The development of the ACOD1 knockout mice (*Acod1*^-/-^), which lacks the ability to produce itaconate, helped elucidate that the endogenous ACOD1/itaconate axis regulates succinate levels, mitochondrial respiration and cytokine production during macrophage activation (11). Particularly, ACOD1 deficiency and a decrease in itaconate production have proven to be deleterious in several acute and chronic inflammatory diseases (12). For example, in response to viral or bacterial infections, *Acod1*^-/-^ mice generally have poorer survival, inflammatory cytokine burst, uncontrolled neutrophil recruitment and immune-mediated tissue damage compared to wild-type (WT) mice (13).

In addition, ACOD1 modulates macrophage polarization in the tumor microenvironment, promoting cancer progression (14). Along with these reports, ACOD1 deficiency has been associated with a worse disease phenotype in a mouse model of psoriasis induced by the TLR7 agonist imiquimod (IMQ) (15). Further, in a bleomycin-induced pulmonary fibrosis model, ACOD1 deficiency promotes persistent fibrosis with increased expression of profibrotic genes (12).

Similar to *Acod1*^-/-^ models, esterified derivatives of itaconate, 4-octyl itaconate (4-OI) and dimethyl itaconate (DI), are commonly used to mimic ACOD1/Itaconate biological effects in vitro and in vivo (16). Indeed, 4-OI has beneficial effects in numerous in vivo animal models (17–19), including our recent findings that this compound attenuates immune dysregulation and organ damage in a mouse model of spontaneous lupus (20). In human studies, treatment of lupus PBMCs with 4-OI decreases the production of pro-inflammatory cytokines (21), while serum itaconate levels are decreased in SLE (22).

Taken together, these reports suggest that the ACOD1/Itaconate pathway could be important in the development and perpetuation of SLE. Therefore, we have now investigated the contribution of endogenous itaconate synthesis to SLE, including its putative role in the development and perpetuation of clinical symptoms and immune dysregulation in the TLR7-induced lupus IMQ model (23). Using a well characterized human SLE cardiovascular cohort, we also investigated whether there is an association between itaconate and subclinical vascular disease in SLE.

## Results

### ACOD1 is induced in murine and human macrophages following TLR7 in vitro stimulation

Givne that, to our knowledge, there were no previous reports on whether TLR7 engagement ( a lupus-relevant pathway) would induce ACOD1, we performed an in vitro time-course on WT and *Acod1*^-/-^ bone marrow-derived macrophages (BMDM) following incubation with IMQ. Western blot analysis showed that ACOD1 protein expression was evident at 12 h of treatment and persisted at 24 h **(Fig. 1a)**. As expected, ACOD1 was not detected in the *Acod1*^-/-^ BMDM. Based on the overall late induction of ACOD1 protein and that type I IFNs are induced by IMQ treatment, we investigated whether the ACOD1 induction relied on type I IFN signaling. To this end, ACOD1 induction was evaluated in BMDM deficient in type I IFN receptor α-1 (*Ifnar1*^-/-^) following in vitro treatment with IMQ for 12 and 24 hours. In the absence of the IFNAR1, ACOD1 induction was significantly decreased, suggesting that ACOD1 expression with TLR7 activation is partially dependent on signaling through IFNAR1 **(Fig. 1b)**. RNA-seq analysis of BMDM treated with IMQ for 12h (GSE250384) showed that *Acod1* is among the 50 differentially expressed genes in response to TLR7 stimulation (**Fig. 1c**). Gene ontology analysis of these IMQ-treated BMDM showed enrichment of pathways associated to cytokines, inflammation, innate immune response, neutrophil degranulation, phagocytosis, and type II IFNs (**Fig. 1d**). Furthermore, the gene expression analysis suggested metabolic rewiring in IMQ-treated BMDM. Specifically, we observed that, under basal conditions, macrophages express higher levels of genes associated with lipid catabolic metabolism, whereas after TLR7 stimulation they switched to pathways of lipid biosynthetic metabolism and express higher levels of genes related to the synthesis of prostaglandins (**Fig. 1e**).

**Fig. 1.**
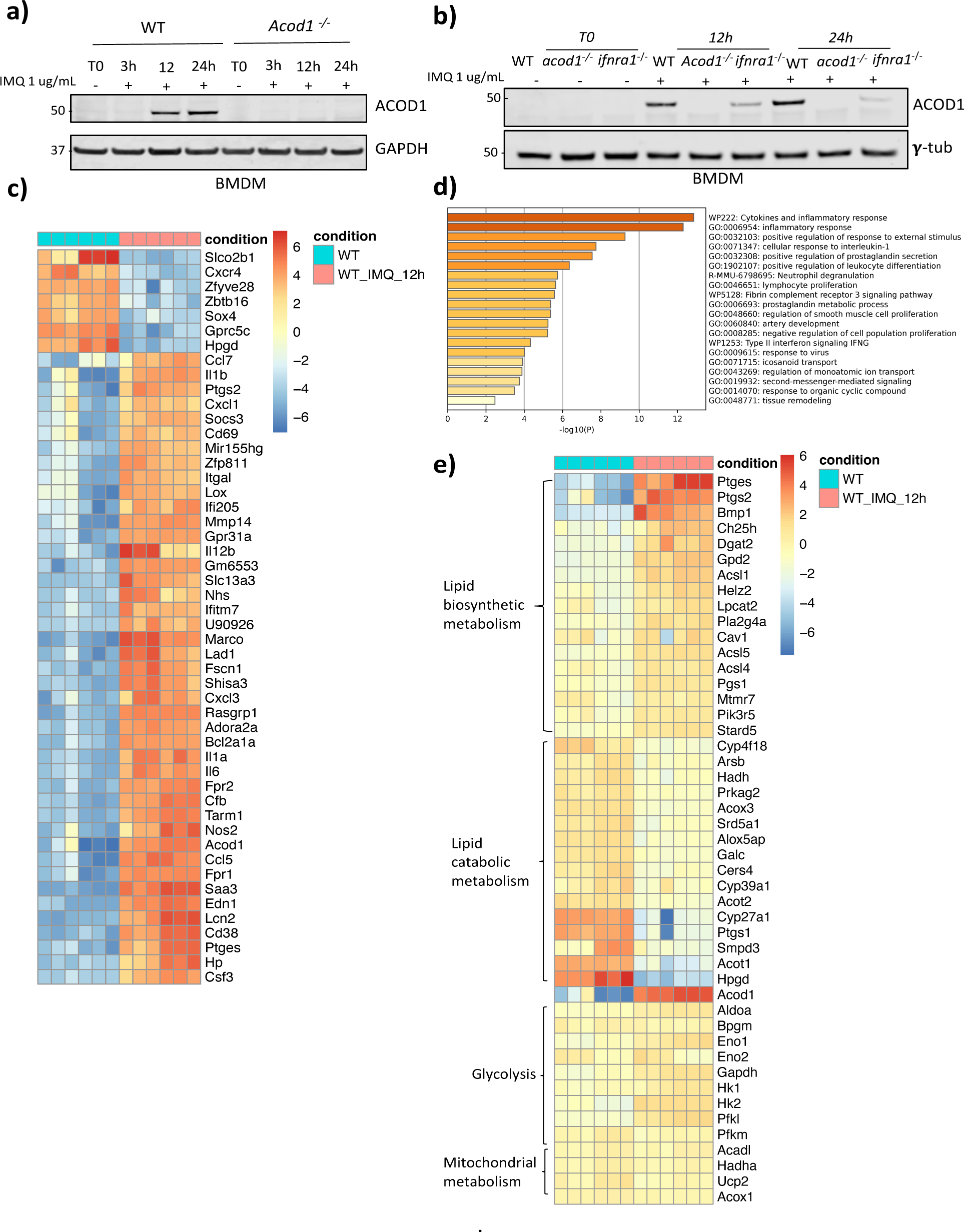
ACOD1 is induced after TLR7 activation in BMDM. **a)** Western blot showing ACOD1 protein expression in WT and *Acod1^-/-^* BMDM treated with imiquimod (1 μg/mL) at 0, 3, 12 and 24 hours. **b)** Western blot showing ACOD1 protein expression evaluated in WT, *Acod1*^-/-,^ and *Ifnar1*^-/-^ BMDM treated *in vitro* with imiquimod (1 ug/mL) for 0, 12 and 24 hours. RNA seq was performed in BMDM treated with IMQ for 12h. Heatmap representation of the genes induced in BMDM after 12h of IMQ stimulation. **d)** Gene ontology analysis of genes differentially expressed between control BMDM and IMQ treated BMDM. **e)** Heatmap representation of the genes associated to metabolic pathways in BMDM after 12h of IMQ stimulation. RNA-seq data was analyzed with DESeq2. The log-transformed DESeq2-normalized read counts were further scaled for each gene individually using the mean expression levels and their corresponding standard deviations to facilitate visualization in a heatmap. Three blots were processed in parallel for a and b, *n = 3*. Six independent mice were chosen from each genotype for c, d, and e. n = *6*.

We proceeded to validate these findings in human healthy control monocyte-derived macrophages. Monocytes were differentiated into M1-like and M2-like using GM-CSF and M-CSF, respectively, then treated with IMQ in vitro for 3 and 12 hours. Similar to BMDM, ACOD1 was induced after 12 hours of IMQ treatment **(Fig. 2a),** but only in the M-CSF (M2)-derived macrophages, perhaps explained by previous observations that GM-CSF-derived macrophages lack TLR7 expression (24) **(Fig. 2b)**. Next, we used specific inhibitors of different signaling pathways reported to be activated downstream of TLR7 stimulation(25). We found that NF-κB, p38, JNK, ERK1/2, JAK-STAT and STING activation are involved in ACOD1 induction after TLR7 activation **(Fig 2c-d)**. In addition, we observed that the induction of ACOD1 mediated by TLR7 activation was decreased after using a histone acetyl transferase II inhibitor, suggesting that epigenetic modifications are also involved in ACOD1 induction after TLR7 activation. Taken together, these results indicate that ACOD1 is induced after TLR7 activation in human and mouse macrophages, in a process that involves type I IFN signaling and canonical pathways of inflammatory cytokine synthesis.

**Fig.2 ACOD1.**
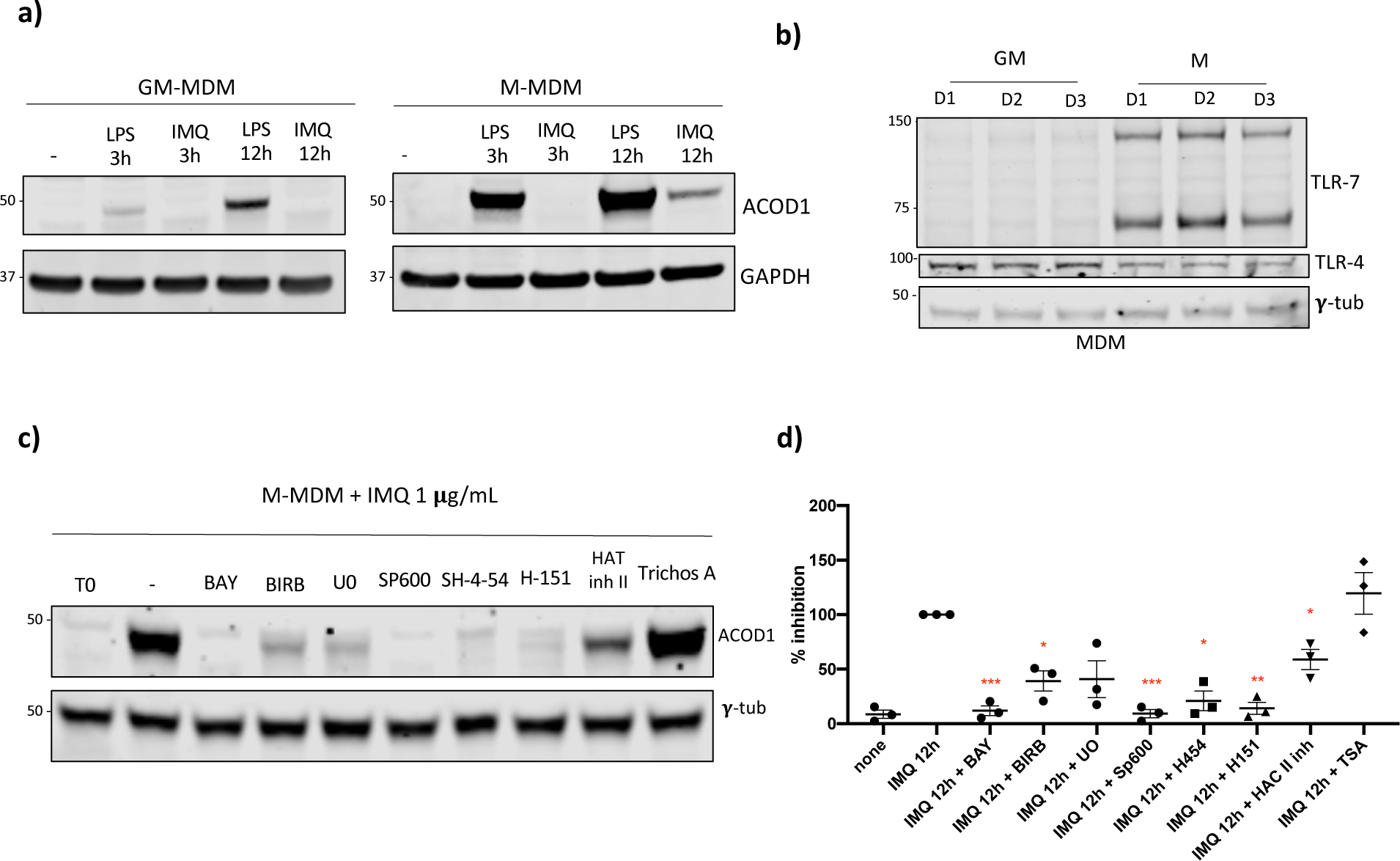
is induced after TLR7 activation in human MDM. **a)** Western blot showing ACOD1 expression in GM and M (GM- and M-CSF, respectively) human MDM (Macrophages Derived from Monocytes) after 12 h of treatment with LPS 100 ng/uL or IMQ 1 μg/mL. **b)** Western blot showing basal expression of TLR4 and TLR7 in GM and M MDM. **c)** Western blot and **d)** densitometry results showing ACOD1 expression in M MDM after TLR7 activation in the presence of different signaling inhibitors. Bay 10 **μ**M (NF-κB), BIRB 0.1 **μ**M (p38), SP600125 30 **μ**M (JNK), U0126 2.5 **μ**M (ERK1/2), SH-4-54 5 **μ**M (STAT3/STAT5), H-151 1 **μ**g/mL (STING), HAT Inh II 5 **μ**M (HAT II), Trichostatin A (TSA) 5 **μ**M (HDAC inhibitor) Graphs show the mean ± SEM, % of expression is relative to the IMQ 12h without inhibitor. *n = 3*, Three blots were processed in parallel for a and c. The statistical analysis was done using Mann-Whitney test, *: p<0.05, **: p<0.01; ***: p<0.005.

### ACOD1 deficient mice develop splenic morphological changes

To further investigate the potential contribution of the endogenous ACOD1/itaconate pathway in lupus autoimmunity, we treated WT and *Acod*1^-/-^ mice epicutaneously with IMQ for 5 weeks. This is a well-established model of induced lupus that is characterized by splenomegaly, type I IFN induction, anemia, renal immune complex deposition, endothelial dysfunction and immune dysregulation (23). For every clinical and most immunologic assessments, we observed that, in the absence of TLR7 stimulation, *Acod1*^-/-^ and WT mice did not show any significant differences.

Mice treated with IMQ (WT and *Acod1*^-/-^) developed an autoimmune phenotype consistent with lupus, and also developed anemia and leukocytosis, as previously described (23) **(Fig. 1S)**. Following IMQ, there were no significant differences in mortality rates comparing WT and *Acod1*^-/^ **(Fig. 3a).** Similarly, no differences between treated *Acod1*^-/^ and WT were detected in serum chemistry (glucose, creatinine, electrolytes). **(Table 1S)**. In contrast, following IMQ treatment, *Acod1*^-/-^ mice displayed lower spleen:body weight ratios compared to WT imiquimod-treated **(Fig. 3b)** Morphologically, IMQ-treated *Acod1*^-/-^ mice displayed marked splenic congestion of the red pulp and atrophy of the white pulp when compared to IMQ-treated WT mice, similar to what has been reported in human SLE (26) **(Fig. 3c)**. Splenocytes from untreated and IMQ-treated WT and *Acod1*^−/−^ mice were analyzed by flow cytometry to quantify immune cell populations. In the IMQ-treated mice, *Acod1*^-/-^ displayed higher percentages of naïve B cells compared to the WT mice, but no changes in the percentage of total B cells, plasma cells, monocytes, DCs, neutrophils **(Fig. 3d)** and T cells **(Fig. 2S).** No differences were found in levels of various type I IFN-regulated genes nor in other genes associated to inflammatory pathways *(Il1b, Il18, Il6, Tlr7 and Tnf*) in splenocytes (**Fig. 3S**) Taken together, results indicate that lack of ACOD1 promotes changes in the splenic architecture but no significant changes in the composition of splenic immune cell types or in regulation of inflammatory genes in a mouse model of lupus.

**Fig. 3.**
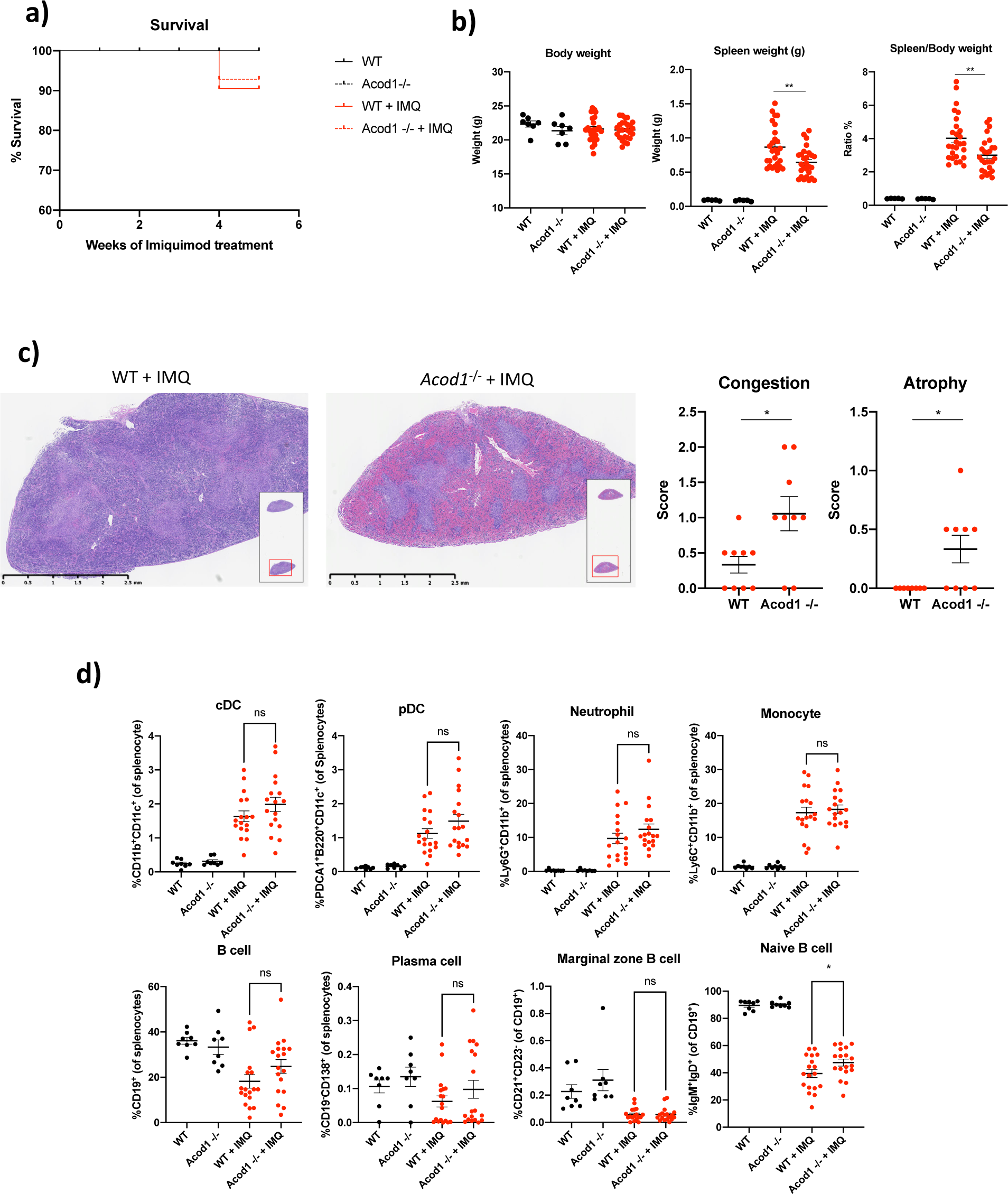
IMQ treated *Acod1*^-/-^ mice display disruptions in splenic architecture and increases in naïve B cells compared to the IMQ-treated WT mice. Wild type (WT) and *Acod1*^-/-^ mice (8–10-week-old female) were treated with 5% imiquimod cream epicutaneously 3 times per week for 5 weeks, to induce a lupus phenotype. **a)** Survival curves for WT and *Acod1*^-/-^ mice during several weeks of imiquimod treatment, *n = 10*. **b)** Spleen weight and spleen:body weight ratio in untreated and IMQ treated WT and *Acod1*^-/-^ mice, *n = 28*. **c)** Spleen H&E staining in formalin-fixed, paraffin-embedded (FFPE) from WT and *Acod1*^-/-^ mice after IMQ treatment, *n = 10*. Pathology scores of spleen congestion and atrophy were as follows: no changes = 0, mild changes = 1, moderate changes = 2, and severe changes = 3. **d)** Splenocytes from untreated and IMQ-treated WT and *Acod1*^−/−^ mice were analyzed by flow cytometry to quantify % of monocytes, DC, neutrophils and B cell populations. Nontreated conditions, *n = 8*, IMQ-treated conditions, n = *18*. Graphs show the mean ± SEM. The statistical analysis was done using Mann-Whitney test, *:p<0.05, **: p<0.01.

### *Acod1*^-/-^ mice have higher levels of serum autoantibodies and kidney immune complex deposition in TLR7-induced lupus

Compared to WT mice, *Acod1*^-/-^ mice exposed to IMQ displayed significantly higher levels of serum anti-dsDNA autoantibodies, while anti-Sm, anti-RNP and total IgG levels were no different from those in IMQ-treated WT mice (**Fig. 4a**). In addition, IMQ-treated *Acod1*^-/-^ mice developed significantly increased glomerular immune complex deposition compared to IMQ-treated WT mice (**Fig. 4b-c**). While this specific mouse model of lupus is not characterized by nephrotic-range proteinuria or end-stage renal disease (27), IMQ-treated *Acod1*^-/-^ mice had increased albumin:creatinine ratios in urine compared to IMQ-treated WT mice (**Fig. 4d**), indicating that renal function was more impaired in *Acod1*^-/-^ mice exposed to IMQ. Consistent with these results, we found that, in kidneys from NZB/W F1 mice, a model of spontaneous severe lupus that we had previously shown improves with exogenous analogs of itaconate, *Acod1* mRNA expression was decreased as disease progressed (10 to 30 week-old mice), suggesting that ACOD1 absence may modulate the development and progression of lupus nephritis (**Fig. 4e**).

**Fig. 4.**
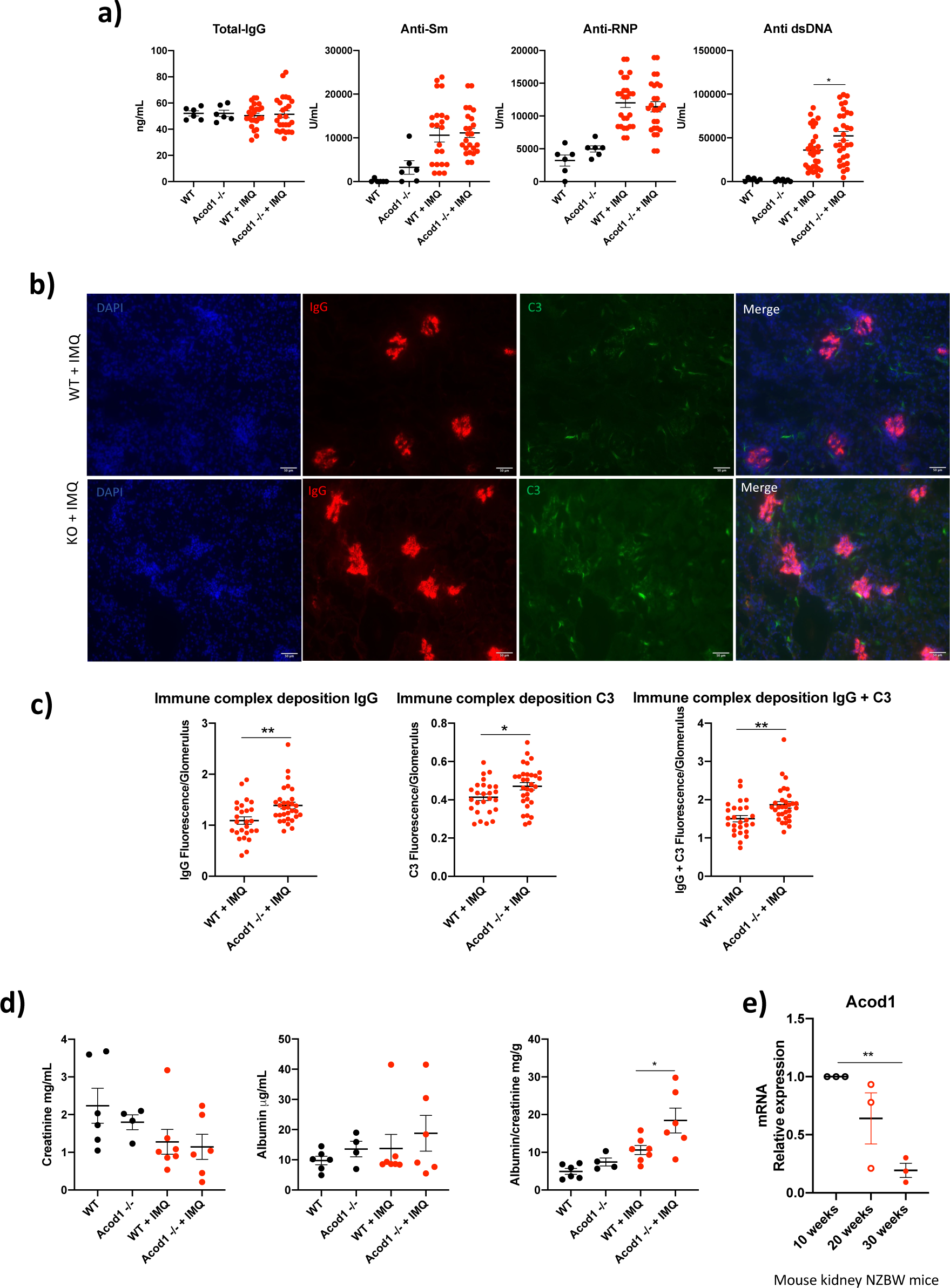
*Acod1*^-/-^ mice treated with IMQ have higher levels of serum anti-ds-DNA, renal immune complex deposition and albuminuria. **a)** Serum levels of total IgG and antibodies against Sm, RNP and dsDNA analyzed by ELISA, untreated conditions, n = *6*. IMQ-treated, *n = 26*. **b)** Representative immunofluorescence photomicrographs displaying renal immune complex deposition. IgG is depicted in red, C3 in green, and nuclei in blue. Original magnification 40x. **c)** Quantification of pixel analysis of glomeruli in 5 different WT or *Acod1*^-/-^ IMQ-treated using ImageJ. n = *5* **d)** Analysis of proteinuria in mice at the time of euthanasia, *n = 7*. Albumin was quantified by ELISA and Creatinine by colorimetric assay; then albumin/creatinine ratios were calculated. **d)** *Acod1* mRNA expression in kidney from spontaneous lupus mice NZBW at 10, 20 and 30 weeks old, *n = 3*. Graphs show the mean ± SEM. Graph in a, c, and e the statistical analysis was done using Mann-Whitney test, *:p<0.05, **: p<0.01. In d, Student’s t test analysis was performed. *:p<0.05.

As endothelial dysfunction and vasculopathy are hallmarks of both murine (28) and human SLE (3, 4), we evaluated endothelium-dependent vasorelaxation in precontracted thoracic aortas following exposure to graded concentrations of acetylcholine. IMQ-treated *Acod1*^-/-^ mice showed similar perturbations in endothelium-dependent vasorelaxation when compared to IMQ-treated WT mice **(Fig. 4S),** suggesting that lack of ACOD1 does not modulate lupus vasculopathy in mice.

As expected, IMQ-treated mice had significant elevation of various pro-inflammatory cytokines compared to the non-treated animals (**Fig. 5S**). Of note, IL-6 serum levels were significantly increased in the IMQ-treated *Acod1*^-/-^ mice compared to the IMQ-treated WT mice, while CCL5 and VEGFA were decreased (**Fig. 5**). Overall, results indicate that ACOD1 modulates pathogenic autoAb synthesis, kidney immune complex deposition and renal injury, and systemic inflammatory cytokine production in lupus-prone mice.

**Fig. 5.**
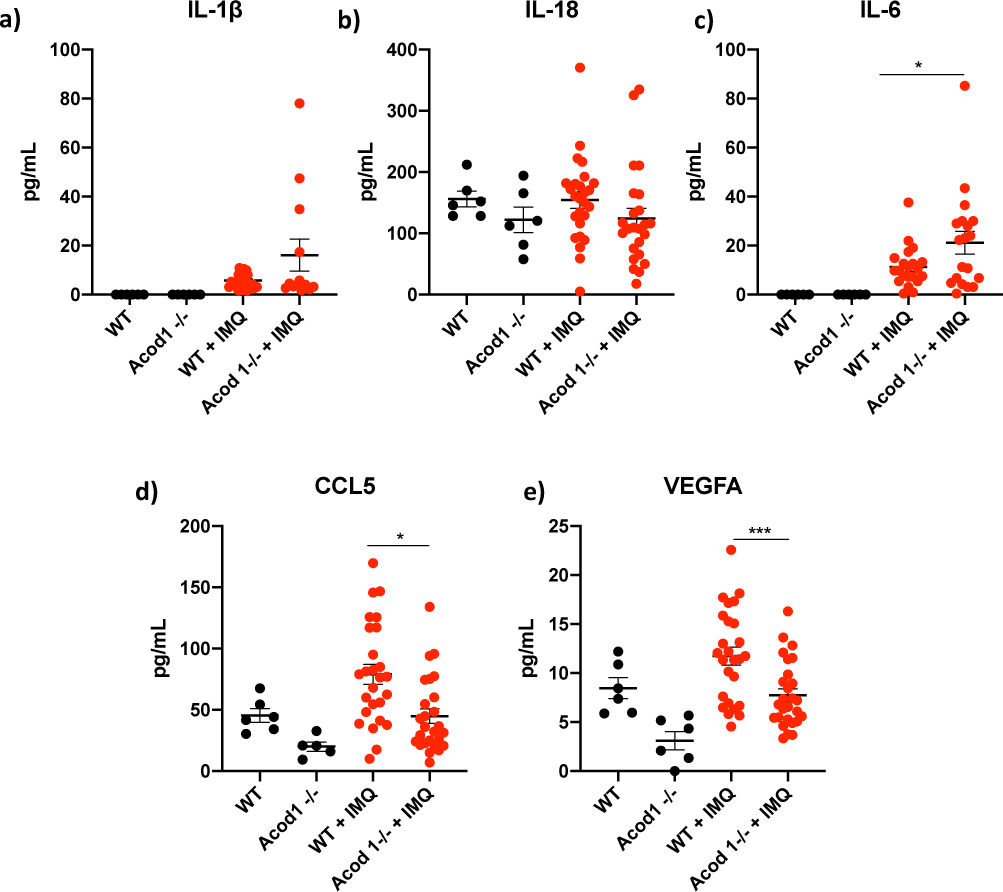
IMQ treated *Acod1*^-/-^ mice have decreased serum levels of CCL5 and VEGFA, while IL-6 serum levels are increased. Detection of various cytokines and chemokines in the serum of WT and *Acod1*^-/-^ mice treated with IMQ. The graphs show the levels for a) IL-1*β*, b) IL-18, c) IL-6, d) CCL5, and e) VEGFA. Untreated conditions, n = *6*, IMQ-treated conditions n = *26.* Graphs show the mean ± SEM. The statistical analysis was done using Student’s t test, *:p<0.05, ***:p<0.005. For CCL5 and VEGFA, IMQ-treated values were adjusted for the untreated values, given that there was a trend for untreated Kos to have lower levels of VEGFA and CCL5.

### *Acod1*^-/-^ modulates myeloid cell responses following TLR stimulation

The ACOD1/itaconate pathway was previously reported to modulate cytokine production in mouse and human macrophages after stimulation with other TLRs or with cytokines (11). To investigate the role of ACOD1 pathway on cytokine/chemokine production following TLR7 stimulation, *Acod1*^-/-^ BMDM were treated with IMQ for 12 hours. qPCR analysis revealed a significant increase in *cxcl2* mRNA levels in IMQ-treated *Acod1*^-/-^ compared to IMQ-treated WT BMDM (**Fig. 6a**), but no significant differences in *il6*, *il1b, ccl2, cxcl9, il18, irf7 and mx1* mRNA levels. Western blot analysis of WT and *Acod1*^-/-^ BMDM treated with IMQ revealed an increase in pro-IL-1β intracellular levels (**Fig. 6b**) and in the secretion of IL-1β and IL-6 (**Fig. 6c**). Consistent with these results, *Acod1*^-/-^ peritoneal mouse macrophages treated for 12 hours in vitro with IMQ showed higher levels of IL-1β and caspase-1 activation (**Fig. 6S**). These results support previous observations that the ACOD1-itaconate pathway regulates IL-1β processing and IL-6 production after activation with other TLRs (TLR4) (11) and suggest that this pathway is implicated in downregulating inflammatory cytokine synthesis triggered by endosomal TLRs.

**Fig. 6.**
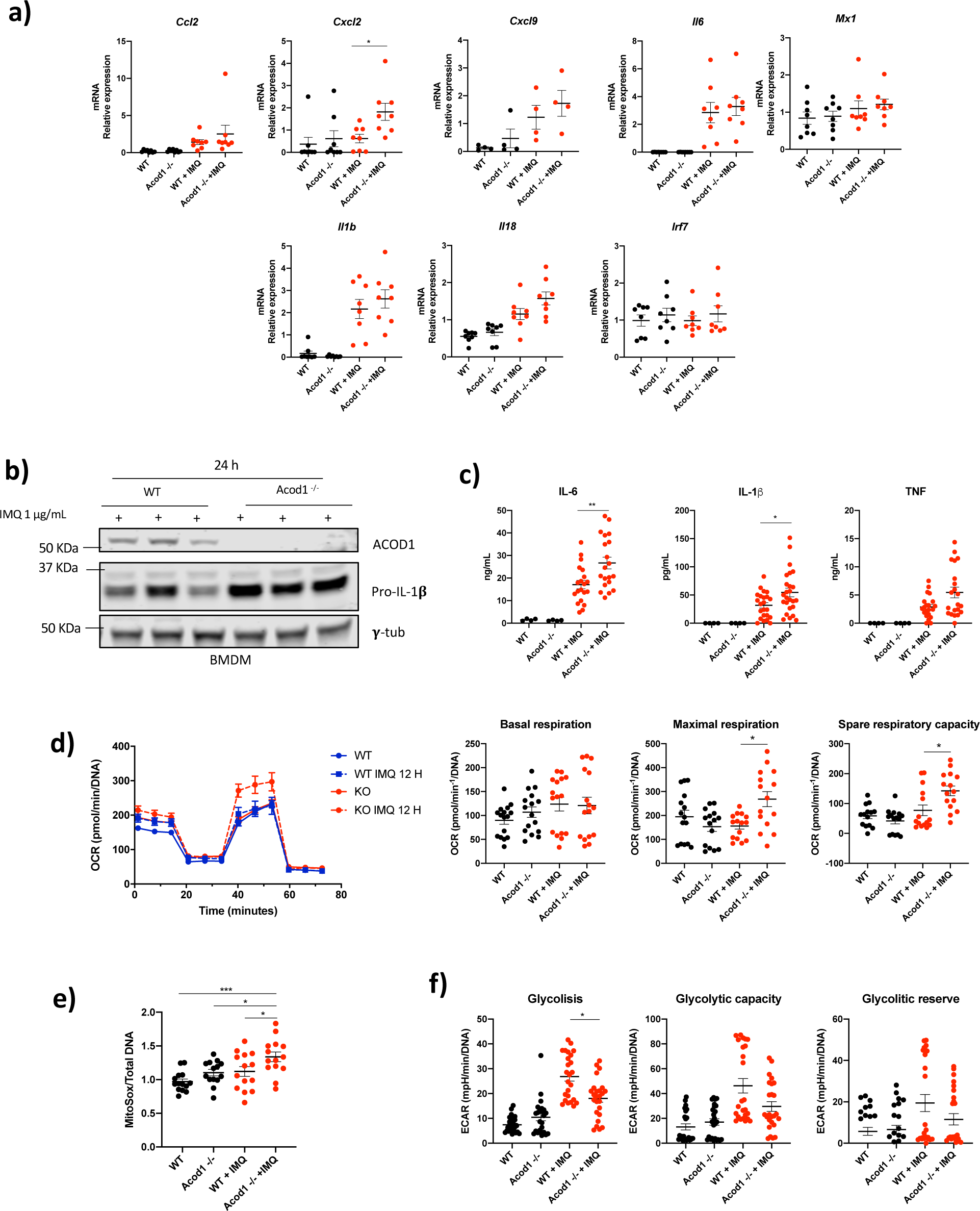
*Acod1*^-/-^ BMDM treated with IMQ display differential levels of proinflammatory cytokines and chemokines and higher mROS production. WT and *Acod1*^-/-^ BMDM were treated with IMQ or left intrerated **a)** after 12 h of stimulation gene expression was measured by qPCR (n = *8*). Data were normalized to untreated mice. Data are represented as mean ± SEM. **b)** Western blot showing pro-IL1 levels in BMDM from three different WT and *Acod1*^-/-^ mice treated with IMQ 1 μg/mL for 24 h. n = *3*. **c)** IL-1*β*, IL-6 and TNF-*α* in the supernatants from WT and *Acod1*^-/-^ BMDM treated with IMQ 1 μg/mL for 24 h. Results are from 4 independent experiments. *n = 20.* d) Seahorse Mitochondrial stress test analysis (n = *15*) and **e)** mitoROS production (mitosox) of WT and *Acod1*^-/-^ BMDM treated or not with IMQ 1 μg/mL for 12h. (n = *15*) **f)** glycolysis stress test analysis of WT and *Acod1*^-/-^ BMDM treated or not with IMQ 1 μg/mL for 24 h (n = *15*). The statistical analysis was done using Mann-Whitney test, *:p<0.05,**:p<0.01.

To further understand the metabolic rewiring induced by ACOD1, we evaluated mitochondrial function in macrophages by measuring the oxygen consumption rate (OCR) using Seahorse analysis. We found that, in the absence of ACOD1, BMDM treated for 12 h with IMQ had higher maximal respiration rate compared to the IMQ-treated WT BMDM (**Fig. 6d**). Consistent with this observation, IMQ-treated *Acod1*^-/-^ BMDM had enhanced mitochondrial ROS (mROS) synthesis compared to the IMQ-treated WT BMDM (**Fig. 6e**). In contrast, glycolysis, was decreased at 24 h of IMQ treatment (**Fig. 6f**). These results support that endogenous itaconate modulates mitochondrial respiration in myeloid cells, as proposed for exogenous itaconate (20).

Increased maximal mitochondrial respiration has been associated with mROS production and oxidative stress during inflammatory conditions. To analyze the effect of ACOD1 absence in other myeloid cells, we tested the ability of BM-derived neutrophils to form NETs, a feature that is dysregulated in murine and human lupus in association with aberrant mROS synthesis (2). Neutrophils from *Acod1*^-/-^ mice treated with IMQ showed no differences in their capacity to form NETs in response to a calcium-ionophore A23187 (**Fig. 7a**) However, *Acod1*^-/-^ BM-derived neutrophils treated in vitro with ionophore A23187 displayed enhanced NET formation compared to the WT neutrophils (**Fig. 7b**). In vivo, IMQ-treated BM neutrophils displayed enhanced mROS production compared to the WT neutrophils (**Fig. 7c**). These data suggest that ACOD1/itaconate pathway attenuates neutrophil activation, NET formation and neutrophil mROS synthesis in response to TLR7 stimulation.

**Fig. 7.**
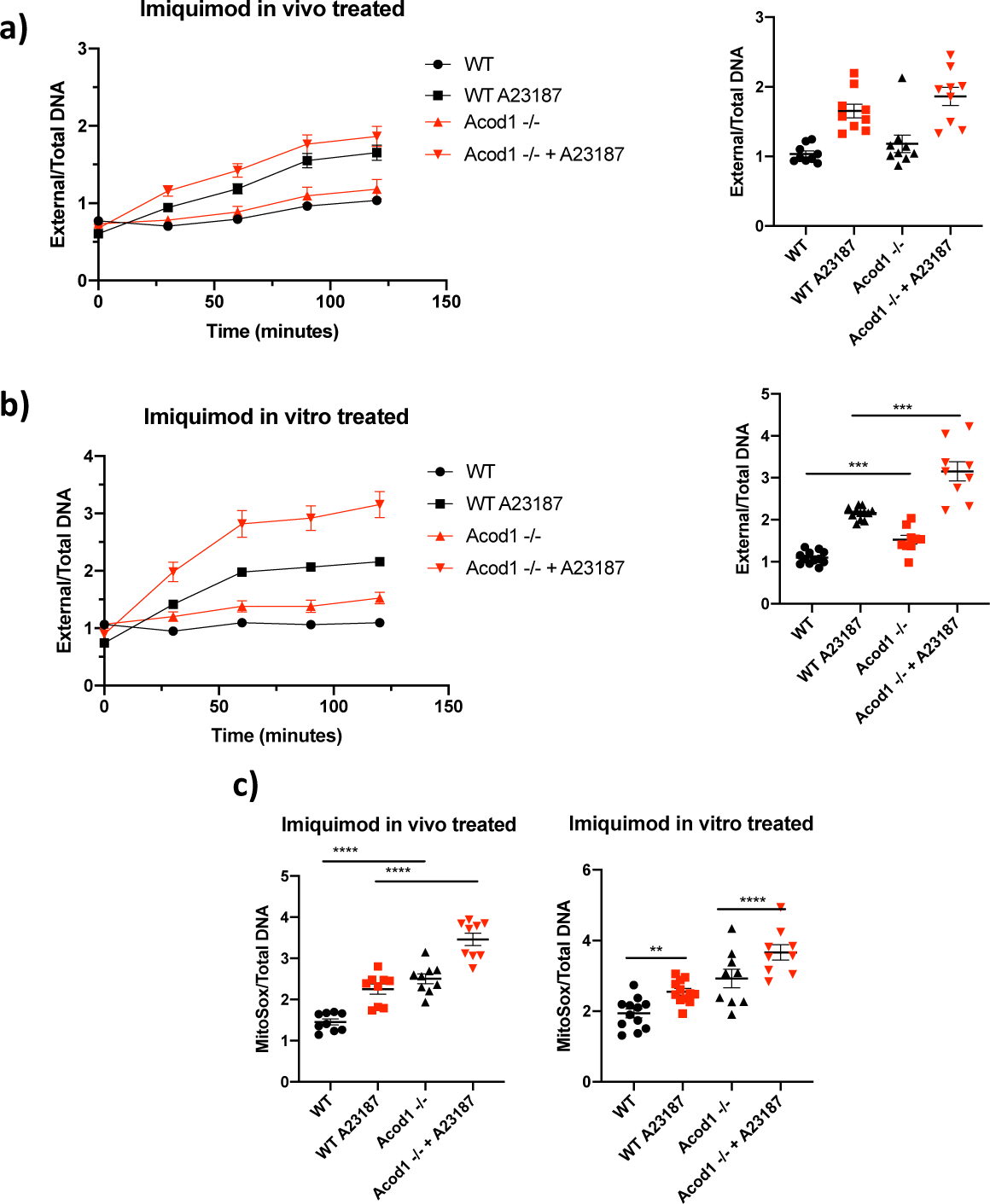
Compared to WT, *Acod1*^-/-^ BM neutrophils have higher NET formation and mitoROS production. **a)** NETs were quantified in BM neutrophils from mice treated with IMQ (in vivo treated) and **b)** BM neutrophils treated in vitro with IMQ 1 μg/mL (in vitro treated), 2 hours post-plating, by Sytox and PicoGreen plate assay to measure external and total DNA, respectively n *= 9*. **c)** Mitochondrial reactive oxygen species (mROS) were quantified 1-hour post-plating, using MitoSox. n *= 9*. Graphs show the mean ± SEM. The statistical analysis was done using Mann-Whitney test, *:p<0.05,**:p<0.01;***:p<0.005;****p<0.001.

### The ACOD1/itaconate pathway is associated with cardiometabolic parameters in patients with SLE

Given the association between ACOD1/itaconate levels and inflammation, we assessed plasma levels of itaconate in SLE patients and found them to be significantly reduced when compared with matched controls (**Fig. 8a**). To investigate the putative clinical relevance of itaconate levels in patients with SLE, correlation analyses were performed with clincail variables. We found a negative association between plasma itaconate levels and lupus disease activity, as measured by the SLE disease activity index (SLEDAI; r=-0.359; p=0.013) and with levels of anti-dsDNA (−0.2, p=0.01). To investigate the putative clinical relevance of itaconate levels with cardiovascular disease in patients with SLE, correlation analyses were performed with various cardiometabolic parameters and assessment of vascular function, vascular wall inflammation and coronary artery plaque. The following parameters were analyzed: arterial stiffness (measured by the cardio-ankle vascular index (CAVI), endothelium-dependent vasorelaxation (measured by reactive hyperemia index (RHI), coronary plaque burden (assessed by coronary CT), vascular wall inflammation (with FDG/PET CT), insulin resistance (assessed by HOMA-IR) and other cardiometabolic parameters, including high density lipoprotein levels and function. The analysis showed that, while itaconate levels positively correlated with arterial stiffness (**Fig. 8b**) and negatively correlated with endothelium-dependent vasorelaxation (**Fig. 8c**), there was a positive association between itaconate levels and higher cholesterol efflux capacity, a marker of atheroprotective HDL function that is typically perturbed in lupus (**Fig. 8e**) and with lower non-calcified coronary plaque burden (NCB) (**Fig. 8d**), body mass index (BMI; **Fig. 8f**), aortic wall inflammation (avg TBR; **Fig. 8g**) and insulin resistance (HOMA-IR (**Fig. 8h**)). These results suggest that dysregulation in itaconate levels in SLE is associated with markers of vascular inflammation and metabolic dysregulation.

**Fig. 8.**
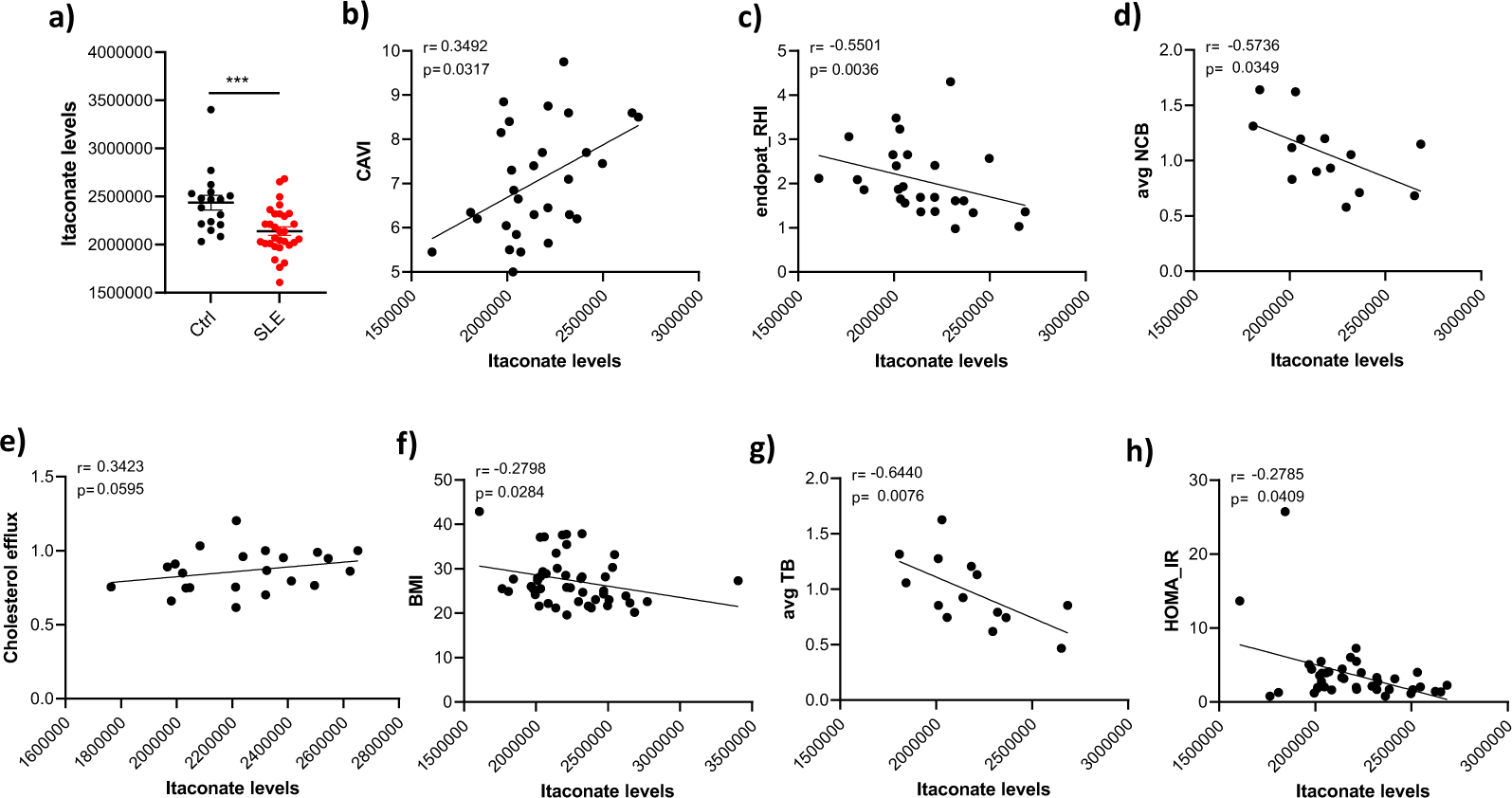
Association between cardiometabolic parameters and itaconate levels in serum in SLE and healthy controls. **SLE** a) Levels of itaconate were measured in plasma samples from SLE patients (n=31) and control (n=17). Results represent mean +/-SEM. Mann Whitney analysis was used. ***p< 0.001. Levels of itaconate measured in SLE plasma were correlated with b) cardio-ankle vascular index (CAVI), c) reactive hyperemia index (RHI), d) non calcified coronary plaque burden (NCB), e) plasma cholesterol efflux capacity (measure of HDL function), f) body mass index (BMI), g) vascular wall inflammation (TB) and h) Insulin resistance (HOMA-IR). Spearman correlation was used.

## Discussion

We previously reported that administration of exogenous itaconate derivatives in the NZB/W F1 mouse model of lupus improves clinical and immunologic features of this disease(20). We have now expanded our investigations on the putative roles of the endogenous ACOD1/ itaconate pathway in mouse and human lupus. Of note, itaconate levels were found to be decreased in SLE compared to healthy individuals, in association with specific clinical, vascular and cardiometabolic parameters, suggesting that dysregulation of this pathway is operational in human disease. We have investigated the role of ACOD1 (the enzyme responsible for itaconate production) in the development of systemic autoimmunity in an induced murine lupus model. We had hypothesized that ACOD1 deficiency would promote immune dysregulation and systemic inflammation in SLE. Consistent with our hypothesis, *Acod1*^−/−^ mice exposed to a TLR7 agonist, a lupus relevant stimulus, displayed disruptions in the splenic architecture, increased serum anti-dsDNA and IL-6, higher kidney immune complex deposition and proteinuria, when compared to the WT IMQ-treated mice. These findings suggest that therapeutic strategies enhancing the ACOD1/itaconate pathway or administration of itaconate derivatives as anti-inflammatory agents could be useful in treating lupus and its associated organ complications.

Various factors have been shown to induce the production of ACOD1/itaconate in macrophages and DCs, including LPS(15, 29), viral and bacterial infections (13) and cytokine stimulation (including type I IFNs) (29). In this regard, we found that ACOD1 is induced in vitro at the mRNA and protein levels in BMDM stimulated with a TLR7 agonist. Further, ACOD1 induction by TLR7 stimulation is partly dependent on type I IFN-dependent signaling, which is consistent with other studies that have shown ACOD1 induction after IFN-β treatment in macrophages.(29, 30).

Recently, the role of macrophages in SLE pathogenesis has gained attention, with a focus on how macrophage polarization may influence disease development. Abnormalities in SLE patients, such as increased levels of type I IFNs, anti-dsDNA, IgG antibodies, NETs, alarmins and mtDNA can skew macrophage polarization towards an M1 phenotype, promoting an hyperinflammatory state and negatively influencing phagocytosis of apoptotic cells (1, 31–33). This M1-like macrophage association and its inflammatory mediators have also been observed in SLE-associated atherosclerosis (34). However, in one of the most severe manifestations of lupus, lupus nephritis (LN), it has been suggested that the most predominant macrophage phenotype is the M2-like phenotype (35). Macrophage polarization experiments showed that ACOD1 is expressed only in human M2-like macrophages following 12 hours of TLR7 stimulation, with no induction observed in M1-like macrophages. It is possible that low serum levels of itaconate in SLE patients could be due, at least in part, to an increase in non-itaconate producing macrophages (M1-like macrophages) versus M2-like. Consistent with this idea, serum levels of the chemokine CCL5, known for inducing macrophage polarization toward an M2-like phenotype (36) were also decreased in the serum of IMQ-treated *Acod1^-/-^* mice compared to the IMQ-treated WT mice. These observations require further investigation to be able to conclude that they indeed play a role in modulation of itaconate levels in SLE. Indeed, we do not rule out that the differential itaconate production in SLE patients could be related to a decrease in ACOD1 catalytic activity (9). Eight active-site residues critical for ACOD1 function have been previously identified (37), with six being classified as very rare and to directly affect active site residues. Along with this idea, severely affected male COVID-19 patients carrying loss-of-function TLR7 variants exhibit decreased levels of ACOD1 mRNA (38). Therefore, understanding the polarization and plasticity of macrophages, together with identification of possible variants in SLE that may be involved in itaconate activity and synthesis would be important in understanding perturbances in immunometabolism, with potential therapeutic implications. This hypothesis requires further investigation.

TLR agonists activate diverse signaling pathways including NF-κB, MAPKs, JAK/STAT, and induction of epigenetic modifications (39). Using different signaling inhibitors, we found that the ability of TLR7 agonists to induce ACOD1 in human macrophages was dependent on NF-κB and MAPK activation. Inhibition of histone acetyl-transferase II also decreased ACOD1 expression, suggesting that epigenetic changes are involved in ACOD1 induction following TLR7 stimulation. Consistent with our observation in BMDM, inhibition of the STING pathway and STAT3/STAT4 also decreased ACOD1 induction. These results are in agreement with previous reports where STING activation after TLR4 engagement induces ACOD1 expression (40).

SLE is a multisystem disease, and its clinical manifestations vary according to the organs involved. We found disruptions in the splenic architecture of the IMQ-treated *Acod1*^-/-^ animals. Given that most immune cell subsets in spleen did not significantly change, it remains to be further determined the mechanisms by which congestion and atrophy are induced, and the functional effects of these abnormalities. Of note, similar findings in the splenic architecture have been reported in human SLE, which may indicate that dysregulation in immunometabolic pathways can contribute to altering the spleen microenvironment. (41, 42). This requires further investigation in future studies.

The effects of ACOD1 absence were also observed in renal function, renal immune complex deposition and in the levels of circulating autoantibodies. Increased immune complex deposition could be the result of higher levels of anti-dsDNA autoantibodies. Indeed, we previously found that 4-octyl itaconate inhibits in vitro B cell proliferation (20); as such, it is possible that endogenous itaconate could have the same inhibitory effect on B cells and plasma cells. It remains to be determined whether increased cell death leading to autoAg generation may be seen when disruptions in endogenous itaconate pathway are operational. However, the observation that NET formation is enhanced in the absence of ACOD1 indicates that the enhanced deposition of immune complexes may be linked to both enhanced autoAb and autoAg formation, at least in part, through neutrophil cell death dysregulation.

The ACOD1/itaconate pathway has been previously described to modulate cytokine production, especially IL-6 and IL1β (15, 29). Consistent with these reports, we found that *Acod1*^-/-^ mice treated with IMQ have higher serum levels of IL-6, results that were consistent with the in vitro macrophage experiments we performed, where IL-6 levels in the supernatant of *Acod1*^-/-^ BMDM treated with IMQ were higher compared to the control BMDM. While IL-1β has been reported to be upregulated in the absence of itaconate (15), our results did not show significant differences in the serum levels of *Acod1*^-/-^ mice treated with IMQ compared to the WT mice IMQ treated. In contrast, in vitro *Acod1*^-/-^ BMDM treated with IMQ have higher levels of IL-1β, which is consistent with other in vitro reports where absence of ACOD1/Itaconate increase levels of IL-1β (15). The modulation of cytokines by the ACOD1/itaconate pathway may be context-dependent, influenced by factors such as the specific cell type, the environment, or the signaling pathways involved. Other factors in the in vivo setting, such as interaction with different cell types, the overall immune response, or compensatory mechanisms could contribute to the observed differences between in vivo and in vitro results.

Neutrophils express ACOD1 and produce itaconate under inflammatory conditions (43). Dysregulated neutrophils have a pathogenic role in SLE (44) and we found that, in the absence of ACOD1, there is enhanced NET formation and enhanced mROS synthesis in TLR7-induced lupus, implicating ROS dysregulation contributing to this phenomenon, as previously described by our group (2). These results are consistent with the recent report that exogenously added 4-OI diminishes formation of NETs by neutrophils of either normal (lean) or obese mice (45).

Noncalcified plaque burden (NCB) significantly increases the risk of plaque vulnerability and rupture as well as the occurrence of acute coronary syndromes (46). Elevated NCB, coupled with intensified arterial wall inflammation, has been linked to a higher prevalence of high-risk plaques in individuals with psoriasis (47). In our study, we found that levels of itaconate in lupus patients negatively correlate with NCB, vascular wall inflammation and insulin resistance, while positively associating with atheroprotective HDL function. These observations implicate alterations in the itaconate pathway potentially playing a role in enhanced vascular risk in SLE, an area that requires further investigation. The findings that vascular stiffness and impaired vascular function associated with itaconate levels appears counterintuitive considering the potentially protective roles in other features of vascular disease, including coronary atherosclerosis and various cardiometabolic parameters. One possibility is differential roles of the ACOD1/itaconate pathway in macrophages and other vascular cells. For example, exogenous analogs of itaconate can inhibit angiogenesis, which could have a deleterious impact in conditions such as SLE, where impaired angiogenesis has been reported (48, 49). It is also possible that the levels of itaconate in association with endothelial dysfunction represent a counter-regulatory function of this pathway to regulate vascular health. Indeed, we found low levels of VEGFA in the serum of IMQ-treated Acod1^-/-^ compared to the IMQ-treated WT mice, which is consistent with recent reports where itaconic acid raises VEGF protein levels in skin and the number of CD31-positive vessels (50). Future studies should explore these different hypotheses.

Limitations of this study include the use of a single mouse model of lupus. However, our previous observations that exogenous analogs of itaconate can improve NZB/W F1, another lupus mouse model, and that levels of ACOD1 decrease in these genetically prone mouse model as nephritis progresses, suggests that these observations are not model-dependent. Nevertheless, future studies should expand our observations into other lupus models as well as other autoimmune disorders. In addition, longitudinal assessments of the cardiometabolic parameters and vascular damage progression in SLE patients will be warranted to further establish the role of itaconate in modulating vascular disease in this disease.

In conclusion, the results from these studies implicate the ACOD1/itaconate pathway in the pathogenesis of SLE and its associated organ damage and indicate that strategies to target aberrant immunometabolism in autoimmunity could have positive therapeutic implications, including a potential role in decreasing organ damage in these devastating conditions.

## Author contribution

EPM and MJK conceived the study, designed experiments, and wrote the manuscript. EPM, SN, LPB, DC, EB, BS, MASB, CCR and AR performed experiments; KJ, FN, SdO, and SB, performed RNA-seq and analysis. ZXY performed histopathologic analysis; YTO contributed patient recruitment and assessment, MD and SH contributed patient evaluation and characterization, MN performed vascular function studies, ZM contributed patient database management, NM contributed study design of vascular studies. EPM analyzed the data and generated the graphs and stats shown in this work.

## Data Availability

RNA sequencing analysis have been uploaded to the GEO with series no. GSE250384. All other data can be requested to the corresponding author.

## Acknowledgments

We thank the Office of Science and Technology, Intramural Research Program, NIAMS/NIH for technical support. We also thank Dr. Kwangwoo Kim (NIAMS/NIH) for helpful discussions. This study was supported by the Intramural Research Program, NIAMS/NIH (ZIA AR041199).

