## Supplementary figure for "The aconitate decarboxylase 1/itaconate pathway modulates immune dysregulation and associates with cardiovascular disease markers in SLE"

**Supplementary information**

**Materials and methods**

**Study approvals.**

All animal study protocols were approved by the animal care and use committee of the NIAMS/NIH, listed on animal study protocol A022-08-04, and in agreement with Animal Research Advisory Committee guidelines (3.18.1). Human studies were approved by the NIAMS/NIDDK IRB (study 94-AR-0066); all subjects signed informed consent and the study was conducted in accordance to Declaration of Helsinki. Patients or the public were not involved in the design, or conduct, or reporting, or dissemination plans of our research.

**Isolation, differentiation, and treatment of BMDM**

Mouse bone marrow cells were flushed out with cold RPMI 1640 medium (Gibco) containing 1% penicillin/streptomycin and were then centrifuged, washed with PBS 1X (Gibco) and filtered through a 70-mm filter mesh. Bone marrow cells were counted and cultured at 2.0 x $10^6$ cells/mL in Dulbecco's modified Eagle's medium (DMEM, Thermo Fisher Scientific), supplemented with 10% fetal bovine serum (FBS, Sigma Aldrich), 1% penicillin/streptomycin, and M-CSF 25 ng/mL (R&D). M-CSF was replaced every 2 days. Six-day differentiated BMDM were left untreated or treated with Imiquimod (IMQ, Invivogen) at 1 μg/mL concentration for the indicated times. Culture supernatants, Trizol and RIPA lysates from stimulated cells were harvested and stored at -80 °C until further evaluation. Six-day differentiated BMDM were left untreated or treated with 1 μg/mL IMQ for the indicated times (1).

**Human monocyte isolation and differentiation into macrophages (MDM)**

Human peripheral blood mononuclear cells were obtained by Ficoll-paque PLUS density gradient (Cytiva) from heparin whole blood of healthy donors using a NIH-approved IRB protocol and following written informed consent. $CD14^+$ monocytes were purified by positive selection using magnetic separation systems (Miltenyi Biotec), as previously reported (2). Monocytes were cultured at $0.5 \times 10^6$ cells/mL for 6 days in RPMI 1640 supplemented with 10 % FBS, 2 mM L-glutamine (Gibco), and 1 % penicillin/streptomycin. Medium was supplemented with 10 ng/mL GM-CSF (Peprotech) or 10 ng/mL M-CSF (R&D Systems) to generate GM-MDMs and M-MDMs, respectively. Cytokines were replaced every-two days. Six-day differentiated macrophages were used for all experiments and, when required, macrophages were treated with LPS (Escherichia coli 0111:B4) at 100 ng/ml or IMQ at 1 μg/mL for the indicated times. Additionally, macrophages were pretreated for 1h with different signaling inhibitors: p38 inhibitor BIRB 0.1μM (Millipore), c-jun N-terminal kinase (JNK) inhibitor SP600125 - 30 μM (Sigma), MAPK kinase 1/2 (MEK 1/2) inhibitor UO126 - 2.5 μM (Millipore), NF-κB inhibitor BAY-117082 - 10 μM (Sigma), STAT3 and STAT5 inhibitor SH-4-54 - 5 μM (Selleckchem), STING inhibitor H-151 - 0.5 μg/mL (Invivogen), The histone deacetylase inhibitor Trichostatin A - 5 μM (Sigma) and a histone Acetyl-transferase II inhibitor - 5 μM (Millipore) were also used.

**Animals and lupus model.**

Female wild type (WT) mice and ACOD1 knockout mice (*Acod1*$^{-/-}$) were purchased from Jackson Laboratory (strains JX5304 and JX29340, respectively). Mice were bred under specific pathogen–free conditions and all treatments and experiments were done in accordance with NIH guidelines (NIAMS protocol no. A019-05-03). To induce a lupus phenotype, WT and *Acod1*$^{-/-}$ mice (8–10-week-old) were treated with 5% IMQ cream (Fougera) epicutaneously, 3 times per week for 5 weeks. Mice were euthanized 48 to 72 h after the final treatment. Blood was collected by terminal cardiac puncture. Spleens and kidneys were harvested for RNA, protein, and histology. Spleens were smashed through a 70 μM sterile filter to obtain single-cell suspensions, and subsequently analyzed by flow cytometry. Red blood cells (RBCs) were lysed using ammonium–chloride– potassium lysing buffer (3-5).

**Complete blood count (CBC).**

CBCs were performed in the NIH Department of Laboratory Medicine.

**Histology**

Spleen samples were fixed in 10% formalin solution. Tissues were sent to Histoserv, Inc for H&E and PAS staining. Histopathology slides were digitally scanned and analyzed by the Pathology Core Facility, NHLBI. Slides were scored based on histopathological criteria as follow: 0, no changes; 1, mild changes, 2, moderate changes and 3 severe changes. Kidney sections were snap frozen and sent to Histoserv, Inc for sectioning.

**Quantification of serum autoantibodies.**

Serum total IgG, anti-Sm, anti-RNP and anti-ds DNA quantification were performed by ELISA using commercially available kits from Alpha Diagnostic International. Mice sera were thawed and clarified by centrifugation at 14,000 rpm 10 min, then diluted 1:100 in low–nonspecific binding buffer. Assays were performed following the manufacturer's instructions (6).

**Evaluation of immune complex deposition in mouse kidney**

For immunofluorescent detection of immune complex deposition, kidneys were perfused with PBS and snap frozen in OCT. Frozen sections were fixed in cold acetone for 10 min and washed with PBS. Slides were blocked with 5% sterile-filtered BSA 1 h at room temperature, then incubated with antibodies diluted in 1% BSA. Samples were washed with PBS and counterstained with 1:10,000 Hoechst. Slides were sealed using Prolong Gold. All images were captured on a Zeiss LSM780 confocal microscope using the same acquisition settings. IgG and C3 were quantified by fluorescence intensity and measured in 10 to 15 independent glomeruli for each mouse using Fiji software. IgG and C3 intensity were subsequently normalized to Hoechst intensity within the same glomerulus to calculate a deposition metric (1).

**Proteinuria evaluation.**

The urine albumin:creatinine ratio was determined using ELISA kits for creatinine and mouse albumin according to instructions of the manufacturer (EthosBiosciences) (6).

**Measurement of serum cytokines, chemokines and growth factors**

Serum obtained from mice at euthanasia was evaluated for the following 48 markers: BAFF, G-CSF (CSF-3), GM-CSF, IFN alpha, IFN gamma, IL-1 alpha, IL-1 beta, IL-2, IL-

3, IL-4, IL-5, IL-6, IL-7, IL-9, IL-10, IL-12p70, IL-13, IL-15/IL-15R, IL-17A (CTLA-8), IL-18, IL-19, IL-22, IL-23, IL-25 (IL-17E), IL-27, IL-28, IL-31, IL-33, LIF, M-CSF, RANKL, TNF alpha; ENA-78 (CXCL5), Eotaxin (CCL11), GRO alpha (CXCL1), IP-10 (CXCL10), MCP-1 (CCL2), MCP-3 (CCL7), MIP-1 alpha (CCL3), MIP-1 beta (CCL4), MIP-2, RANTES (CCL5); Betacellulin (BTC), Leptin, VEGFA. Soluble receptors: IL-2R, IL-7R alpha, IL-33R (ST2). The Invitrogen™ ProcartaPlex™ Mouse 48-plex panel (Thermo Fisher Scientific) was used to quantify these molecules. Mice sera were thawed and clarified by centrifugation at 14,000 rpm 10 min and processed according to instructions of the manufacturer (7).

**RNA sequencing**

RNA was isolated from BMDM exposed or not, to IMQ for 12 hours. Tri Reagent (Sigma-Aldrich) was used followed by column-based purification with RNA Clean & Concentrator-5 Kit (Zymo Research). NEBNext Poly(A) mRNA Magnetic Isolation kit (New England BioLabs) was used to generate cDNA libraries. Picogreen dsDNA quantitation assay (Thermo Fisher Scientific) was used to measure the cDNA concentration; cDNA was diluted to 3 nM, cDNA libraries were pooled together, and RNASeq data were generated with an Illumina Novaseq6000 system (8).

**Bulk RNA-seq analysis**

Sequencing data were generated with an Illumina Novaseq6000 system. The FastQ files were produced by processing raw sequencing data using bcl2fastq (v2.20.0). Subsequently, paired end sequence reads were processed with Partek Flow version

10.0.22.1204 (Partek Inc, https://www.partek.com/partek-flow/). The trimmed reads were mapped onto mouse genome mm10 using STAR 2.7.8a as implemented in Partek Flow with default settings. Quantification was done using the Partek Flow "quantify to annotation model (Partek E/M)" and the Ensembl transcript release 101. The gene counts were used for differential gene expression assessed using DESeq2 (v1.42.0) (9). Heatmaps were generated using the R package pheatmap (v1.0.12). GO term analysis was performed using Metascape (10). RNAseq data were deposited in the Gene Expression Omnibus (http://www.ncbi.nlm.nih.gov/geo/) under accession GSE250384.

**Seahorse analysis of BMDM**

Seahorse analysis was performed as previously described (11). The following reagents were used: glucose, oligomycin, carbonyl cyanide-4-(trifluoro-methoxy) phenylhydrazone, 2-deoxy-D-glucose, rotenone, antimycin A, sodium pyruvate (all from Sigma Aldrich), L-glutamine (Agilent), XF calibrant (pH 7.4; Agilent), and XF RPMI medium (pH 7.4; Agilent). Seahorse plates and cartridges were from Agilent. BMDM were plated on Seahorse culture plates (100,000 cells/well) in XF RPMI medium. Analysis was performed at 37°C with no CO2, using an XF-96e analyzer according to instructions of the manufacturer (Agilent). Mitochondrial stress test assay was performed using Seahorse XF RPMI medium with 25 mM glucose, 1 mM sodium pyruvate, and 2 mM L-glutamine. For mitochondrial stress tests, cells were treated serially with oligomycin (5 μM), FCCP (1 μM), rotenone (100 nM), and antimycin A (1 uM), and oxygen consumption rates were quantified. For glycolysis stress tests, same number of cells were suspended in Seahorse XF RPMI medium with 2 mM L-glutamine; cells were treated serially with glucose (25 mM), oligomycin (5 μM), and 2-DG (100 mM), and extracellular acidification rates were measured over time. Cell numbers at assay completion were normalized to DNA content using CyQuant dye (Thermo Fisher Scientific). Wave, Excel, and GraphPad Prism software were used to analyze and graph the data (11).

**Quantitative real-time PCR**

RNA from macrophages was obtained, and gene expression was evaluated by qRT-PCR. Total RNA was extracted using Tri Reagent (Sigma-Aldrich) and retrotranscribed with the Iscript™ Reverse Transcription Supermix cDNA Synthesis kit (Bio-rad). For qRT-PCR, mRNA levels of selected genes were detected using TaqMan® _Gene Expression Assays (Applied Biosystem Inc.) and results were normalized relative to the expression of TATA-box binding protein (*TBP*) and glyceraldehyde-3-phosphate dehydrogenase (*GAPDH*) mRNA and expressed using the Δ Δ cycle threshold method. Gene expression was quantified using a CFX96 Real-Time PCR System (Bio-rad) (2).

**Western blot**

Protein lysates were subjected to SDS-PAGE and transferred onto polyvinylidene difluoride membranes (Biorad). Unspecific binding was blocked with 10 % BSA (Sigma Aldrich) for 1h. Membranes were then incubated with antibodies against proteins of interest and incubated with the corresponding secondary IRDye 800 CW antibodies (Li-cor). Bands were detected using the Odyssey CLx Instrument (2).

**Cytokine determination in cell supernatant**

Culture supernatants from cells stimulated with TLR agonists were harvested, and release of TNF, IL-1 β, IL-6 was quantified with ELISA kits (BD Biosciences).

**Quantification of NETs and mROS.**

Isolation of mouse bone marrow neutrophils, quantification of NET formation and mROS were performed as previously described (1). Briefly, BM neutrophils were purified with Percoll gradient. Cells were seeded in 96-well plates (200,000 cells/100 µl/well in triplicates for each dye) and allowed to form NETs in the presence of Sytox (to quantify extracellular DNA; 1 µM final concentration), Quant-It PicoGreen (to quantify total DNA; stock solution diluted 1:250), and MitoSOX (to quantify mROS; final concentration 200 ng/ml). All dyes were from Thermo Fisher Scientific. At baseline, 1 h, and 2 h, fluorescence was measured for PicoGreen (485 of 520 nm), MitoSOX (510 of 580 nm) and Sytox (485 of 520 nm), respectively, using a FluoStar Omega BMG Labtech plate reader. Cells without dye were used as blanks.

**Patient recruitment**

Healthy adults without any prior clinical cardiovascular disease and adult patients that fulfilled revised criteria for SLE (12) were recruited from the NIH Healthy volunteer cohort and the NIAMS Lupus clinic or NIAMS Community Health Center (13). Briefly, SLE subjects with an estimated GFR < 60 ml/min/1.73 m2 BSA were excluded from coronary CT angiogram (CCTA). Pregnant or lactating women, as well as individuals with active infections, were not eligible for participation. Controls were excluded if they had any concurrent medical issues or were taking medications that could potentially interfere with study results. Following enrollment, controls were further excluded if they exhibited HDL <40, BMI >35, diagnosed hypertension, and/or dyslipidemia. All participants underwent a thorough physical examination and provided fasting blood and urine samples before undergoing vascular testing. Laboratory parameters including fasting blood glucose and lipid panel, white blood count with differential, and systemic inflammatory markers were quantified in the clinical laboratory at the NIH: HOMA-IR was calculated as (glucose [nmol/ml] + insulin [μIU/ml])/22.5.

**Vascular Functions tests**

Lupus patients and healthy had been recruited to a previously described cardiovascular cohort at NIH (13) and underwent various vascular function, coronary artery plaque and vascular wall inflammation, cholesterol efflux capacity assessment and measurement of insulin resistance using HOMA-IR. Briefly, subjects were instructed to abstain from smoking, caffeinated beverages, and fasting for at least 6 hours before testing, withholding medications on the test morning. Testing occurred in a temperature-controlled room with subjects in a supine position. CAVI was measured using VaSera-1500, with cuffs and EKG electrodes placed on arms and ankles. Results were automatically calculated with VaSera VS-1000 software. Peripheral Arterial Tonometry (PAT): Microvascular endothelial function was assessed using EndoPAT 2000. Finger probes were applied bilaterally, and a BP cuff was placed on one arm. With the other arm serving as the control arm. PAT was continuously measured for 20 minutes, and — in between, for 5 minutes — BP cuff was inflated to supra systolic pressure in the test arm. After occlusion dilatation, RH was captured by EndoPAT as an increase in the PAT signal amplitude and compared with the control arm.

A postocclusion/preocclusion ratio was calculated by EndoPAT software, providing a RHI.

CCTA: Performed with a 320-detector row scanner, coronary plaque in major arteries was assessed using QAngio CT. Total (TB), calcified (CB), and noncalcified burden (NCB) indices were calculated. The imaging process included localizer images, coronary calcium score, contrast timing images, and contrast-enhanced images.

**Metabolite Sample Preparation**

Plasma samples were thawed on ice and precipitated using an equal volume of ice-cold methanol. An equal volume of chloroform was added to each sample. Samples were agitated for 30 minutes at 4 $^{o}$C and subsequently centrifuged at 16k g for 20 min to induce layering. An aliquot of the top (aqueous) and bottom (organic) layer were collected. The aqueous layer was diluted 5x in 1:1 methanol:water for LCMS injection. For all LCMS methods LCMS grade solvents were used (14).

**Measurement of itaconate in serum from SLE patients**

Measurement of itaconate serum samples from healthy volunteers and SLE patients was evaluate by Liquid Chromatography Tandem Mass Spectrometry as previously described (15). Briefly, aqueous metabolites were analyzed using a combination of a previously established ion-pairing method operating exclusively in negative ionization mode with modification and a pentafluorophenylpropyl (F5) column method operating exclusively in positive ionization mode (McCloskey, Schwarz, Groveman). Ion-pairing injections were separated using a Sciex ExionLC AC system and measured using a Sciex 5500 QTRAP mass spectrometer. Quality control samples were injected regularly to monitor for signal stability. For negative mode analysis, peaks were resolved with on a Waters Atlantis T3 column (100 Å, 3 μm, 3 mm X 100 mm) with a gradient from 5 mM tributylamine, 5 mM acetic acid in 2% isopropanol, 5% methanol, 93% water (v/v) to 100% isopropanol over 15 minutes. F5 injections were separated on a Phenomenex Kinetex F5 column (100 Å, 2.6 μm, 2.1 mm X 100 mm) with a binary gradient from 100% water with 0.1 % formic acid to 95 % acetonitrile with 0.1 % formic acid over 5 minutes at a flow rate of 0.3 mL/min. In either method, metabolite was identified using at least two distinct multiple reaction monitoring (MRM) signals and a defined retention time. All signals were integrated using SciexOS 3.0. Signals with greater than 50% missing values were discarded and remaining missing values were replaced with the lowest registered signal value. All signals with a QC coefficient of variance greater than 30% were discarded. Metabolites with multiple MRMs were quantified with the higher signal to noise MRM. Filtered datasets were total sum normalized prior to analysis and stitched together using common signals for arginine and glutamine. Single and multi-variate analysis was performed in MarkerView Software 1.3.1 (14, 16).

**Correlation of serum itaconate levels with vascular function tests.**

Vascular function tests were performed as previously described (13). Cardio-ankle vascular index (CAVI), Endopath_RHI, non-calcified burden (NCB), Cholesterol efflux, Body mass index (BMI), vascular inflammation (TB) and Insulin resistance (HOMA IR) were calculated, and each of them correlated to serum itaconate levels.

**Statistics.**

Data were plotted using GraphPad Prism. Statistical tests for group comparisons were performed as appropriate. For endothelium vasorelaxation assays, two-way analysis of variance with Tukey's correction for multiple comparisons was used. For all other analyses, Mann-Whitney, Kruskal-Wallis analysis or Student's t test were performed to determine significant differences. P values less than or equal to 0.05 were considered significant.

**Data availability:** RNA sequencing analysis have been uploaded to the GEO with series no. GSE250384. All other data can be requested to the corresponding author.

**Materials and methods references.**

### Supplementary Results

### Supplementary Figure 1

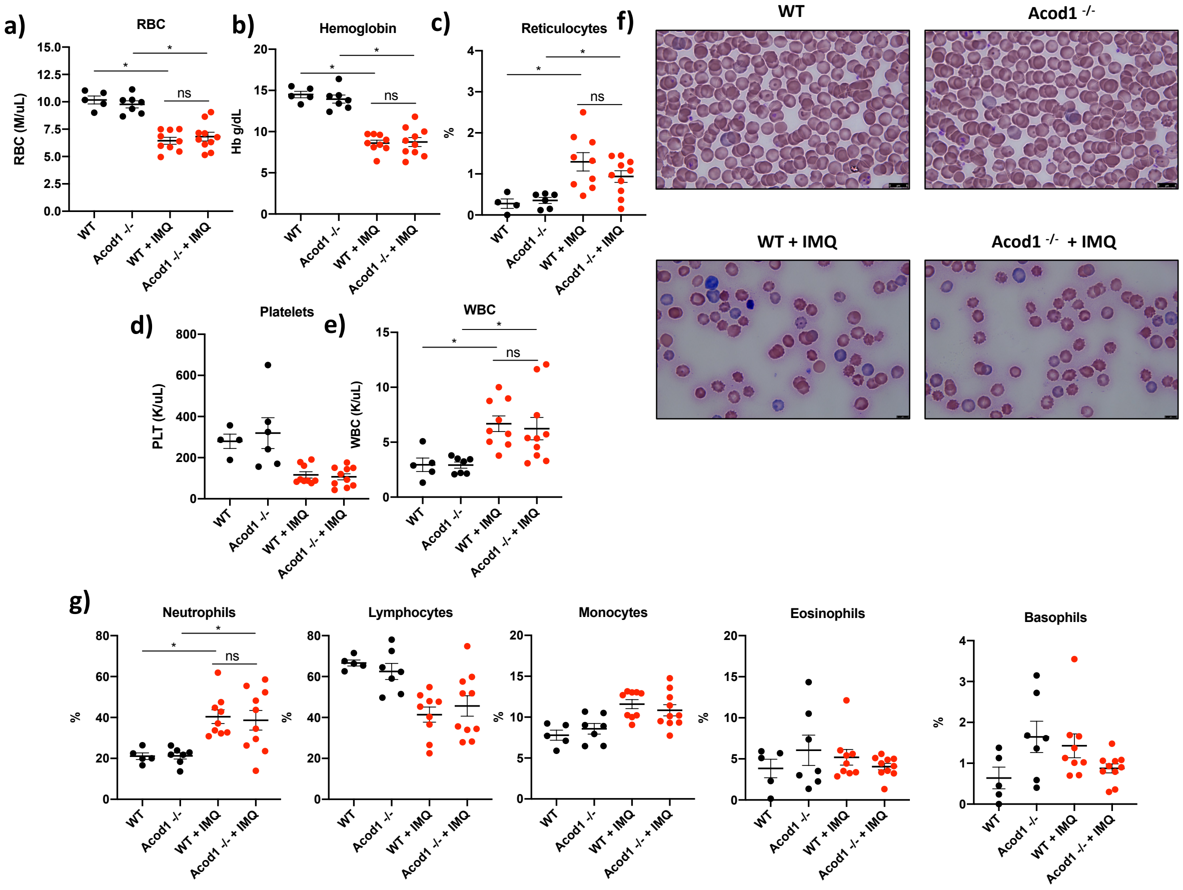


**Supplementary Figure 1. CBC and Wright staining of peripheral blood in untreated or IMQ-treated WT and *Acod1*$^{-/-}$ mice.** a) Red blood cell count (M/µL), b) Hemoglobin (g/dL), c) Percentage of reticulocytes, d) Platelets (K/µL), e) White blood cells count (K/µL), f) red blood cell morphology by Wright staining, and g) Total number of neutrophils, lymphocytes, monocytes, eosinophils and basophils. Untreated WT and *Acod1*$^{-/-}$ conditions, n = *6.* IMQ-treated WT and *Acod1*$^{-/-}$ conditions, n = *10.* Graphs show the mean ± SEM. The statistical analysis was done using Student's t test,*:$p<0.05$.

**Supplementary Table 1. Complete metabolic panel on serum from WT and *Acod1*$^{-/-}$ mice untreated or treated with IMQ for 5 weeks.**

| Biochemistry parameters in WT vs KO Acod1 imiquimod treated animals | | | | | | |
|---|---|---|---|---|---|---|
| **TEST** | Units | WT | Acod1 -/- | WT + IMQ | Acod1 -/- + IMQ | Normal values |
| **ALB** | g/dL | 4.88 ± 0.1 | 4.71 ± 0.2 | 2.62 ± 0.2 | 2.76 ± 0.1 | 2.5 - 4.8 |
| **ALP** | U/L | 92.17 ± 12.5 | 108.29 ± 10.5 | 62.11 ± 4.8 | 64.4 ± 4.6 | 62 - 209 |
| **ALT** | U/L | 72 ± 25.7 | 96.33 ± 15.4 | 64.56 ± 7.7 | 55.8 ± 7.5 | 28 - 132 |
| **AMY** | U/L | 1175.16 ± 294.8 | 896 ± 38.6 | 900.88 ± 36.0 | 869.9 ± 59.8 | 1691 - 3615 |
| **TBIL** | mg/dL | 0.36 ± 0.1 | 0.34 ± 0.0 | 0.27 ± 0.0 | 0.31 ± 0.0 | 0.1 - 0.9 |
| **BUN** | mg/dL | 17.33 ± 0.9 | 16.57 ± 0.9 | 16.44 ± 0.8 | 14.2 ± 0.6 | 18 - 29 |
| **Ca** | mg/dL | 12.51 ± 0.3 | 12.41 ± 0.2 | 11.53 ± 0.2 | 11.4 ± 0.8 | 5.9 - 9.4 |
| **PHOS** | mg/dL | 14.23 ± 0.9 | 16.2 ± 2.1 | 9.22 ± 0.3 | 9.85 ± 0.7 | 6.1 - 10.1 |
| **CRE** | mg/dL | 0.2 ± 0.0 | 0.25 ± 0.0 | 0.21 ± 0.0 | 0.24 ± 0.0 | 0.2 - 0.8 |
| **GLU** | mg/dL | 335.83 ± 33.2 | 365.71 ± 29.0 | 214.33 ± 8.5 | 216 ± 6.7 | 90 - 192 |
| **Na+** | MMOL/L | 158.5 ± 3.5 | 159.5 ± 1.5 | 154.87 ± 1.1 | 154.22 ± 0.3 | 124 - 174 |
| **K+** | MMOL/L | 12.68 ± 1.0 | 12.21 ± 0.8 | 10.22 ± 0.1 | 9.99 ± 0.2 | 4.6 - 8.0 |
| **TP** | g/dL | 5.95 ± 0.1 | 5.84 ± 0.2 | 4.37 ± 0.2 | 4.84 ± 0.2 | 3.6 - 6.6 |
| **GLOB** | g/dL | 1.06 ± 0.1 | 1.11 ± 0.1 | 1.7 ± 0.1 | 1.96 ± 0.1 | 0.0 - 0.6 |
| | | | | | | n = 10, ± st error |

The results show the mean ± standard deviation. No significant changes were observed.

### Supplementary Figure 2

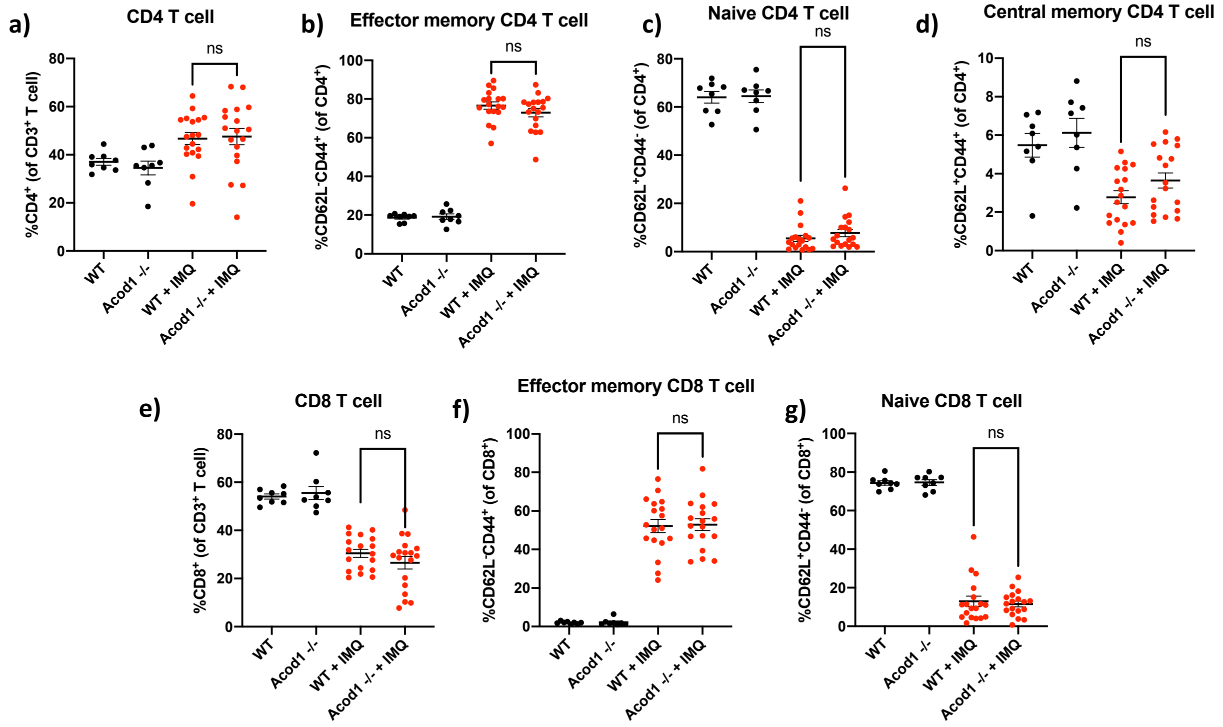


**Supplementary Figure 2 Flow cytometric analysis of T cell subsets in spleen from untreated and IMQ-treated WT and *Acod1*$^{-/-}$ mice:** a) CD4$^{+}$ T cells, b) effector memory CD4$^{+}$ T cells, c) naïve CD4$^{+}$ T cells, d) central memory CD4$^{+}$ T cells, e) CD8$^{+}$ T cells, f) effector memory CD8$^{+}$ T cells, and g) naïve CD8$^{+}$ T cells. Untreated conditions, $n = 8$, IMQ-treated conditions, n = *18.* Bars show the mean ± SEM. The statistical analysis was done using Mann-Whitney test, *:p<0.05,**:p<0.01;***:p<0.005;****p<0.001.

### Supplementary Figure 3

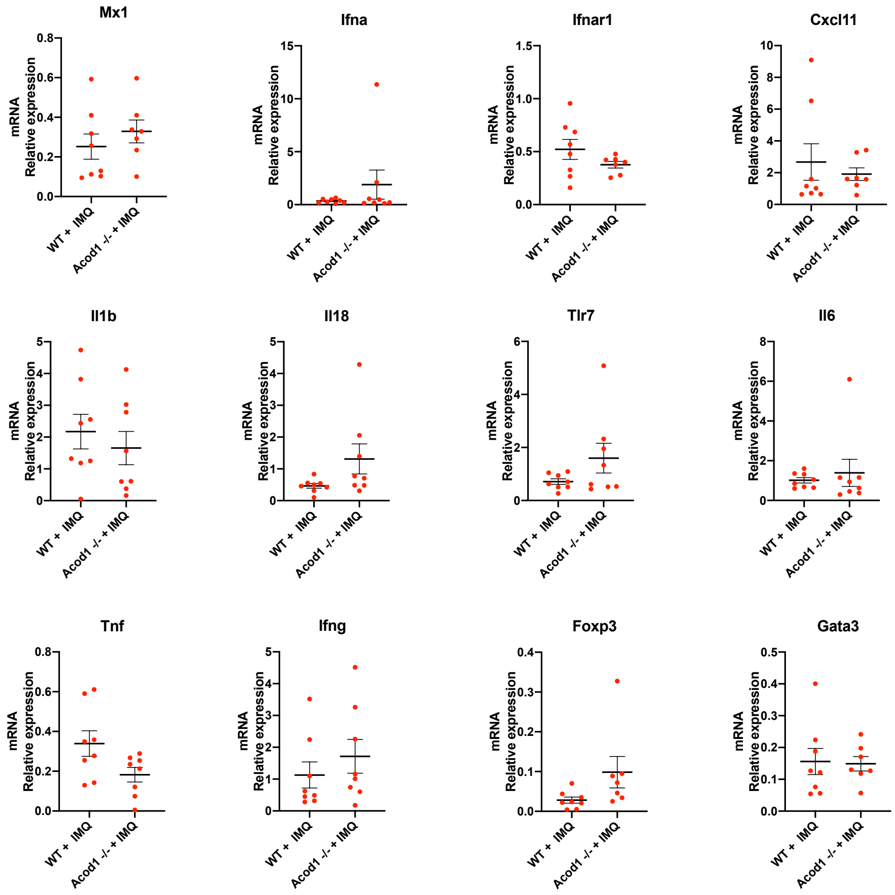


**Supplementary Figure 3. No differential gene expression was found in splenocytes from IMQ-treated WT mice compared to IMQ-treated *Acod1*$^{-/-}$ mice.** Gene expression in splenocytes was analyzed by quantitative real-time polymerase chain reaction (qRT-PCR) normalized against GAPDH (housekeeping gene). n = *8.* Bars show the mean ± SEM. The statistical analysis was done using Mann-Whitney test, no significant changes were found.

**Supplementary Figure 4**

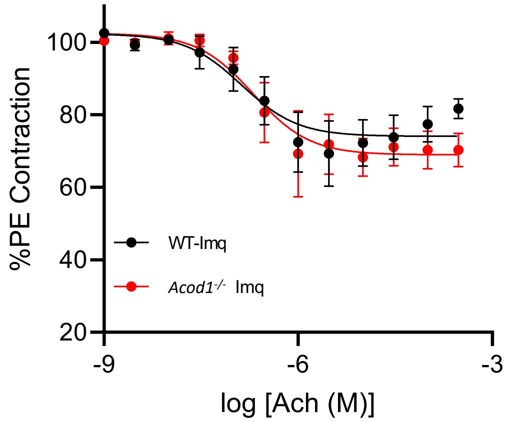


**Supplementary Figure 4 ACOD1 does not modulate endothelium-dependent vasorelaxation.** Endothelium-dependent vasorelaxation was assessed by myograph evaluation of aortic rings from WT and *Acod1*$^{-/-}$ IMQ treated mice in response to acetylcholine after full contraction induced by phenylephrine. Shown in the graph are the averages ± SEM. n = *5*. The statistical analysis was done using Mann-Whitney test. No significant difference was found.

### Supplementary Figure 5

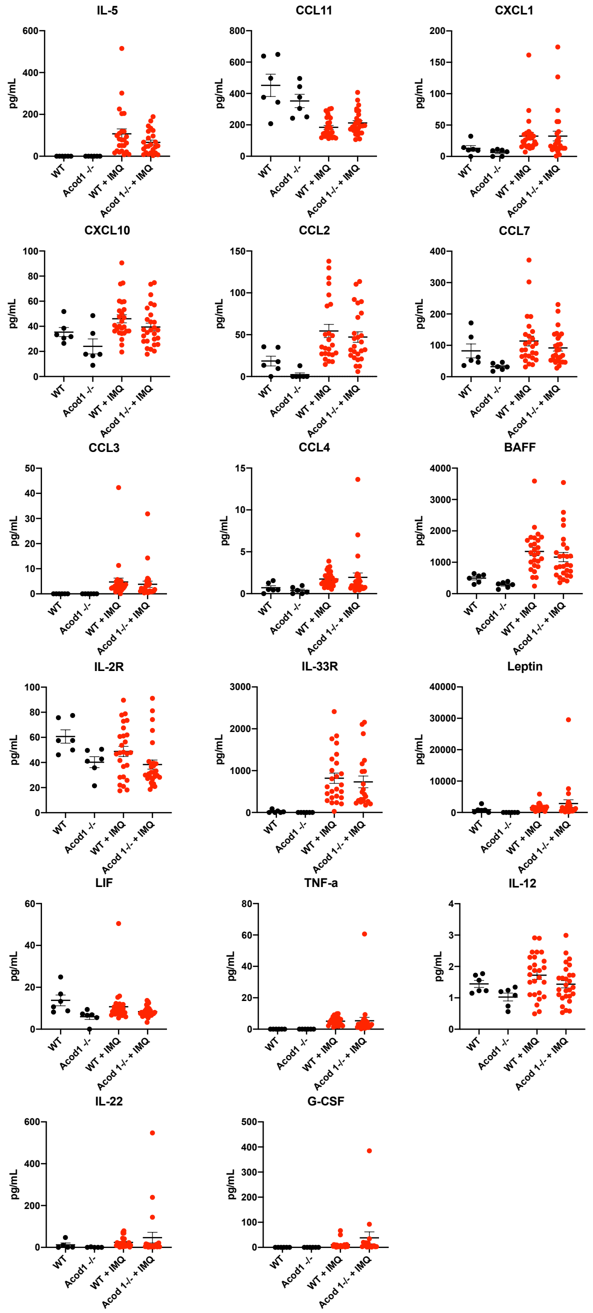


**Supplementary Fig. 5 Cytokines and chemokines that were not affected by lack of ACOD1.** Detection of various cytokines and chemokines in the serum of WT and *Acod1*$^{-/-}$ mice treated with IMQ. Bars show the mean ± SEM. Untreated conditions, n = *6*, IMQ-treated conditions n = *26.* The statistical analysis was done using Student's t test.

### Supplementary Figure 6

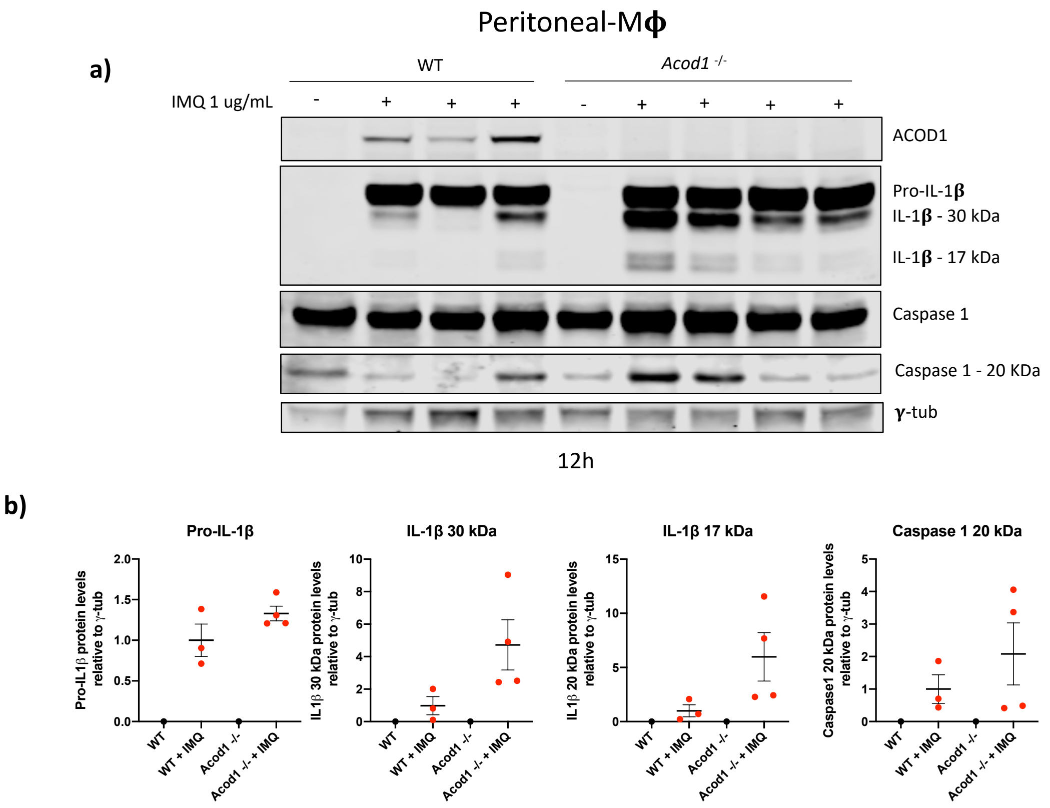


**Supplementary Figure 6 ACOD1 deficient peritoneal macrophages have increased levels of IL-1β. a)** Western blot showing pro-IL1 and caspase 1 levels in peritoneal macrophages from non-treated (WT mice, n = *1* and *Acod1*$^{-/-}$ mice, n = *1*) and IMQ-treated (WT mice, n = *3* and *Acod1*$^{-/-}$ mice, n = *4*) in vitro with IMQ 1 µg/mL for 12 h. **b)** Densitometry analysis of indicated proteins. The statistical analysis was done using Mann-Whitney test.

**Supplementary Figure 7**

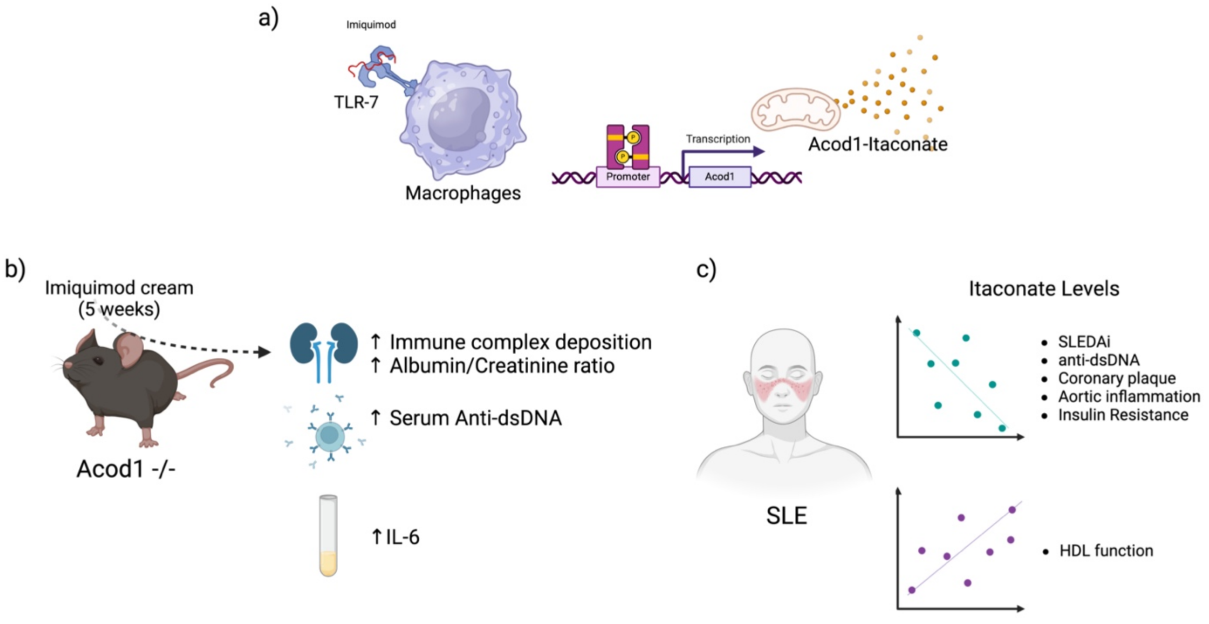


**Supplementary Figure. 7 Graphical Abstract. a)** After TLR7 activation, ACOD1 expression is induced in bone marrow derived macrophages. This induction is partially dependent on type I interferon (IFN) signaling. Lack of ACOD1 increases *cxcl2 mRNA,* IL-6 and IL-1$\beta$ proteins levels. **b)** In an induced lupus model ACOD1-deficiency induces innate and adaptive immune dysregulation. Evidenced by splenic red pulp congestion and white pulp atrophy, increased renal immune complex deposition and proteinuria, elevations in serum autoantibodies and inflammatory cytokines, with decreases in CCL5 and VEGFA. **c)** While in SLE patients, itaconate levels associates with cardiometabolic parameters. ACOD1 may play an important role in regulating the immune system and preventing tissue damage in autoimmunity.

Created with BioRender.com
